# Deciphering the functional impact of Alzheimer’s Disease-associated variants in resting and proinflammatory immune cells

**DOI:** 10.1101/2024.09.13.24313654

**Authors:** Marielle L. Bond, Ivana Y. Quiroga-Barber, Susan D’Costa, Yijia Wu, Jessica L. Bell, Jessica C. McAfee, Nicole E. Kramer, Sool Lee, Mary Patrucco, Douglas H. Phanstiel, Hyejung Won

**Author notes:** Corresponding authors: Douglas H. Phanstiel, Hyejung Won. These authors have contributed equally to this work (Marielle L. Bond, Ivana Y. Quiroga-Barber).

## Abstract

Genome-wide association studies have identified loci associated with Alzheimer’s Disease (AD), but identifying the exact causal variants and genes at each locus is challenging due to linkage disequilibrium and their largely non-coding nature. To address this, we performed a massively parallel reporter assay of 3,576 AD-associated variants in THP-1 macrophages in both resting and proinflammatory states and identified 47 expression-modulating variants (emVars). To understand the endogenous chromatin context of emVars, we built an activity-by-contact model using epigenomic maps of macrophage inflammation and inferred condition-specific enhancer-promoter pairs. Intersection of emVars with enhancer-promoter pairs and microglia expression quantitative trait loci allowed us to connect 39 emVars to 76 putative AD risk genes enriched for AD-associated molecular signatures. Overall, systematic characterization of AD-associated variants enhances our understanding of the regulatory mechanisms underlying AD pathogenesis.

---

Alzheimer’s disease (AD) is a progressive neurodegenerative disease that impacts over 26 million people around the world^1^. Despite its global burden, treatment options are limited, in large part because molecular mechanisms underlying AD remain unclear. Common variation is a major source of risk for AD, explaining 33% of total phenotypic variance^2^. The most recent genome-wide association study (GWAS) identified 75 genomic regions associated with AD^3^. The critical next step is to distill biological mechanisms of AD risk from common variant associations.

Understanding the functional impact of common variation is challenging because most GWAS-identified genetic variants reside in non-coding genomic regions with poorly understood functionality^4,5^. Causal variants are thought to alter the regulatory activity of non-coding loci. However, a GWAS locus typically contains dozens of non-coding variants, each requiring high-throughput functional validation. Once causal variants are identified, pinpointing their target genes is also challenging, because non-coding loci can alter transcription of genes millions of base pairs away via chromatin loops^6–9^. Importantly, these variant-gene relationships are dynamically regulated in a cell type- and context-specific fashion^10,11,12–15^, highlighting the importance of understanding the functional impact of risk variants in the relevant cellular context.

Multiple lines of evidence suggest that AD risk variants promote disease by altering gene regulation in immune cells^16,17^. Previous studies have shown that genetic risk factors for AD are enriched in open chromatin regions in macrophages and microglia^13,18^. Moreover, peripheral monocytes and macrophages have been shown to infiltrate AD brains^19^, and complex crosstalk between peripheral and central immune cells has been proposed to contribute to neuroinflammation and disease progression in AD^20,21^. Finally, AD brains have been found to harbor specific subclasses of microglia known as disease associated microglia (DAMs) that exhibit a distinct proinflammatory expression profile and correlate with overall amyloid level and neurofibrillary tangle pathology^22–24,12–15^. Therefore, it is important to study the regulatory impact of AD risk variants in immune cells such as macrophages and microglia, not only under resting conditions but also in a proinflammatory state, to better model the pathogenic process of AD development.

Here we employed a massively parallel reporter assay (MPRA), a high-throughput technology for quantifying regulatory activity of genetic variants^25–29^, on 3,576 AD risk variants spanning 25 GWAS loci^30^ in resting and LPS+IN-Fγ-treated macrophages, which we refer to as “proinflammatory macrophages.” Empirical characterization via MPRA identified 47 putative causal AD risk variants across 12 loci that show both enhancer and allelic activity. To understand the regulatory impact of these variants, we leveraged gene regulatory networks using RNA-seq, H3K27ac ChIP-seq, ATAC-seq, and Hi-C datasets in resting and proinflammatory macrophages^31^. Together, we mapped empirically validated variants to 76 putative causal AD risk genes, which are enriched for aging-related molecular signatures and DAM signatures. Finally, we quantified the impact of a putative causal AD risk variant on transcription factor (TF) binding using electrophoretic mobility shift assays (EMSAs). These findings identify a set of putative causal variants and risk genes for AD, providing targets for further research and therapeutic development.

## Results

### MPRA identifies shared and condition-specific regulatory elements

To quantify the regulatory activity of AD-associated GWAS variants, we designed an MPRA library to test all variants with nominal significance (p < 1 × 10^-5^) within the GWAS loci described by Jansen et al^30^. We filtered out those that overlapped restriction enzyme sites used for molecular cloning and those longer than 1 bp in length, resulting in 3,498 variants (**Sup Figure 1a**). We synthesized 150-bp sequences flanking these variants (hereafter referred to as AD-associated genetic element), for both risk and protective alleles, and cloned them upstream of a minimal promoter driving GFP and a 20 bp barcode (**Figure 1a**). We also included CMV and EF1 promoters as positive controls and 198 scrambled 150-bp sequences as negative controls (**Sup Figure 1b**). We packaged this MPRA library into lentivirus and introduced it into THP-1 monocytes that were differentiated into macrophages (**Sup Table 1**). Macrophages were used because their open chromatin exhibits the same level of enrichment for AD GWAS variants as microglia^18^. To model an AD-related pathogenic condition, we tested cells in both resting conditions and after stimulation with LPS and IFNγ, which were previously shown to induce a proinflammatory transcriptional profile similar to that observed in disease-associated microglia (DAM)^32^ (**Figure 1a**). We isolated RNA barcodes from macrophages and DNA barcodes from the lentivirus, and quantified the transcriptional activity of each AD-associated genetic element by its RNA/DNA ratio^29^ (**Sup Table 2**). Our library contained a median of 107 barcodes per variant to mitigate potential barcode effects on transcription (**Sup Figure 1c**). We obtained a total of 9 biological replicates for resting macrophages and 6 biological replicates for proinflammatory macrophages resulting in a median Pearson correlation coefficient of 0.84 and 0.63 for resting and proinflammatory macrophages, respectively (**Sup Figure 1d-e**).

**Fig. 1.**
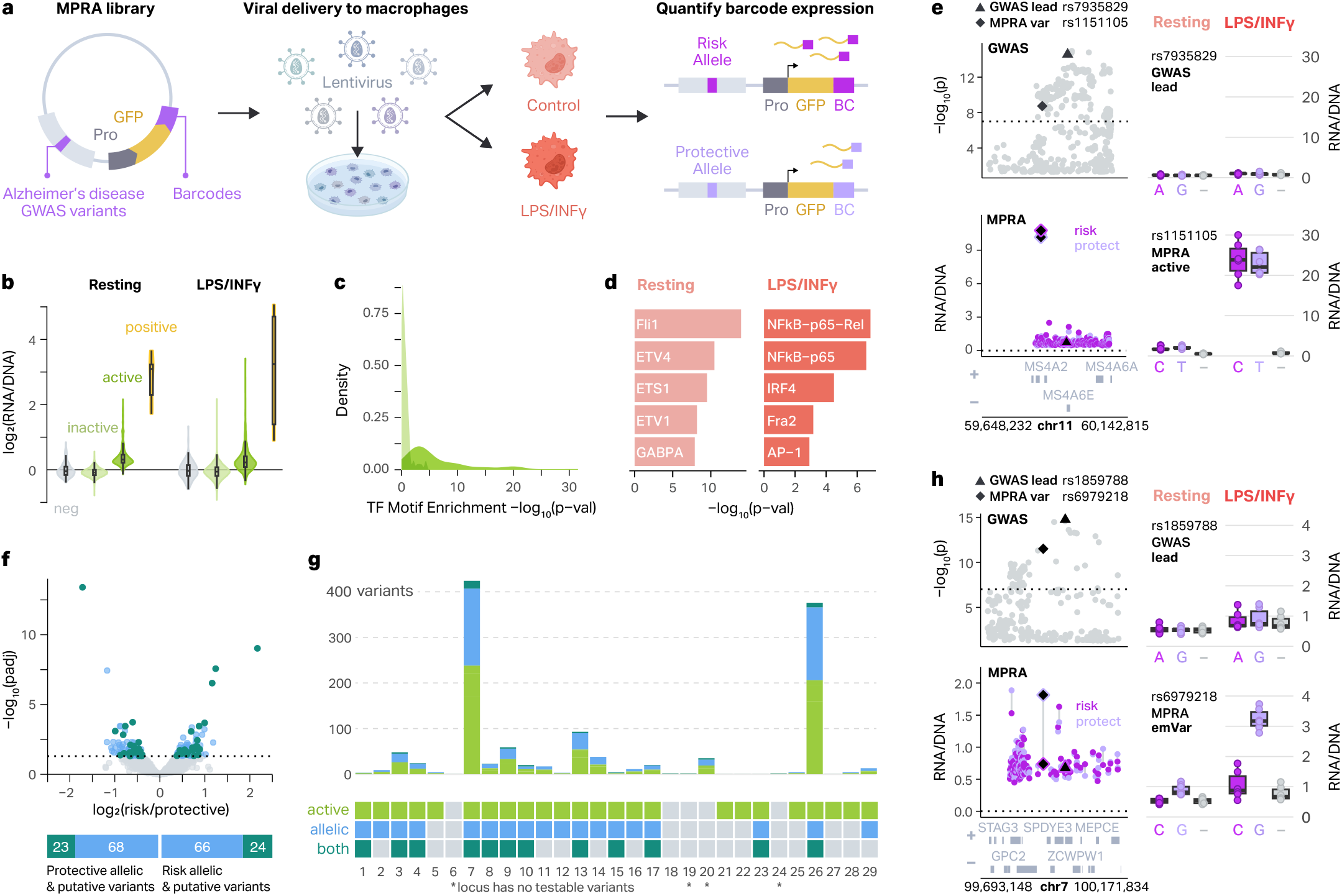
MPRA on AD-associated genetic variants identifies functional regulatory variants. **a**, Schematic of the experimental design. The MPRA library includes 3,576 AD-associated variants upstream of a minimal promoter (Pro), GFP reporter gene, and 20 bp barcode. The library was packaged into lentivirus and introduced into THP-1 monocytes, which were differentiated into macrophages. Barcode expression from risk and protective alleles was compared in resting and LPS+INFγ-treated macrophages to define MPRA-active elements and MPRA-allelic variants. **b**, MPRA activity is shown as log_2_(RNA/DNA) barcode ratio for negative controls (neg), inactive MPRA elements, active MPRA elements, and positive controls (pos) for resting (left) and LPS+INFγ-treated (right) macrophages. Two-sided Wilcoxon rank-sum tests were conducted in both conditions between the log_2_(RNA/DNA) ratios for MPRA-active versus MPRA-inactive elements (p < 2.2 x 10^−16^ resting; p < 2.2 x 10^−16^ LPS+INFγ). Violin plots represent the overall distribution of the data. Box plots represent the median and 25th to 75th quartile (interquartile range, IQR) and whiskers extend to the most extreme non-outliers. **c**, Density plot showing the distribution of –log_10_ p-values of enrichment of TF motifs in MPRA-active and MPRA-inactive elements (Two-sided Wilcoxon rank-sum test, p = 1.0 x 10^−7^). **d**, Top 5 TF motifs enriched in MPRA-active elements in resting (left) and LPS+INFγ-treated (right) macrophages compared to a background set including all inactive elements. **e**, Example of context-specific MPRA activity at the *MS4A6A* locus. Manhattan plot showing GWAS –log_10_ p-values (top left) and MPRA activity for each pair of alleles for each variant (bottom left). GWAS lead variant is represented by a triangle and MPRA-active variant is represented by a diamond. Boxplots showing the activity of each allele for the GWAS lead variant (top right) and MPRA-active variant (bottom right) in both conditions versus negative control (–), where the boxplots show the median and IQR with whiskers extending to the most extreme non-outliers. **f**, Volcano plot showing allelic-regulatory activity of 3,498 AD-associated variants (top, limma-based mpralm, FDR < 0.05). Bar plots showing which MPRA-allelic variants (green) are also MPRA-active elements (teal), or emVars. **g**, Barplot showing the number of MPRA-active and/or MPRA-allelic variants at each locus (top). Legend showing which loci have at least one MPRA-active and/ or MPRA-allelic variant (bottom). Asterisk represents loci which have no testable variants. **h**, Example of a context-specific emVar at the *ZCWPW1* locus. Manhattan plot showing GWAS p-values (top left) and MPRA activity (bottom left) for each variant. GWAS lead variant is represented by a triangle and MPRA active variant is represented by a diamond. Box plots show the activity of each allele for the GWAS lead variant (top right) and MPRA emVar (bottom right) in both conditions versus negative control (–). The box plots show the median and IQR with whiskers extending to the most extreme non-outliers.

First, we identified AD-associated genetic elements with significantly higher regulatory activity compared to scrambled controls. As expected, sequences encoding CMV and EF1 promoters exhibited high regulatory activity in both conditions (median log_2_(RNA/DNA) ratio 3.09 for resting macrophages and 3.24 for proinflammatory macrophages; **Figure 1b, Sup Fig 2a**). In addition to these positive controls, 553 AD-associated genetic elements exhibited higher regulatory activity compared to scrambled controls (linear mixed effect model for the MPRA activity, FDR < 0.05) which we refer to as “MPRA-active elements’’ (**Figure 1b, Sup Fig 2b, Sup Table 3**). MPRA-active elements were enriched for TF binding motifs compared to MPRA-inactive elements (two-sided Wilcoxon rank-sum test, p = 1.0 × 10^−7^; **Fig 1c, Sup Fig 2c**) and included binding motifs for TFs with known roles in macrophage biology including ELF1, SP1, and AP-1.

Since enhancers are often regulated in a condition-specific manner^33–36^, we next characterized the impact of proinflammatory stimuli on regulatory activity of MPRA-active elements by comparing resting to proinflammatory macrophages. Of MPRA-active elements, 212 (38%) showed differential transcriptional activity between two conditions (linear mixed effect model for interaction effects, FDR < 0.05). Resting condition-specific MPRA-active elements were enriched for the erythroblast transformation specific (ETS) TF family, including FLI1, ETV4, and ETS1. In contrast, proinflammatory condition-specific MPRA-active elements were enriched for TFs involved in proinflammatory activation including NFKB, IRF, and AP-1 family (**Figure 1d**). These data support the validity of the MPRA-measured transcriptional activity and demonstrate that the regulatory activity of the AD-associated genetic elements is sensitive to the biological context. An example of MPRA-active elements with condition-specific enhancer activity is at the *MS4A6A* locus. In this locus, we detected an MPRA-active variant rs1151105 that is only transcriptionally active in proinflammatory macrophages, yet there is no difference in the activity of alleles in either context (**Figure 1e**). Notably, rs1151105 has much higher enhancer activity than the GWAS lead variant rs7935829.

### MPRA quantifies the regulatory impact of AD-associated gene variants

Next, we surveyed the allelic activity of AD-associated variants by comparing the regulatory activity between protective and risk alleles. We identified 181 AD GWAS variants that exhibited significant differences in regulatory activity between alleles (limma-based mpralm, FDR < 0.05), which we refer to as “MPRA-allelic variants^29^” (**Figure 1f, Sup Fig 2b, Sup Table 4**). MPRA-allelic variants were also more enriched for TFs than MPRA-non allelic variants, albeit to a lesser degree than MPRA-active elements. These allelic variants would be difficult to predict from GWAS data alone as our MPRA-active elements and MPRA-allelic variants only included 2 lead GWAS variants, and were on average 92.4 kb away from the lead variants (**Sup Fig 2d**). Moreover, neither MPRA-active elements nor MPRA-allelic variants exhibited differences in GWAS p-values or effect size when compared to MPRA-inactive elements and MPRA-non allelic variants respectively (**Sup Fig 2e-h**).

Most of the GWAS loci (25 out of 29) had MPRA testable variants. Out of the 25 loci investigated, 24 harbored at least one MPRA-active element or MPRA-allelic variant. Previous studies have stated the importance of identifying “expression-modulating variants,” or emVars^25^. These are variants that are both MPRA-active and MPRA-allelic by our definitions, and are more likely to be the true regulatory variants. In total, we identified 47 emVars across 12 GWAS loci. We hypothesize that these emVars are potential causal variants driving genetic association with AD (**Figure 1g, Sup Fig 2b, Sup Table 4**).

Given the level of context-specificity detected in MPRA-active elements, we performed differential analysis of MPRA-allelic variants between resting and proinflammatory macrophages. Notably, 140 out of 181 (77.3%) MPRA-allelic variants displayed statistically significant interaction between allelic activity and condition (FDR < 0.05, **Sup Table 4**). A striking example of this is at the *ZCPW1* locus. The lead variant of this locus, rs1859788, was neither active nor allelic in either condition. However, we identified an MPRA-allelic variant, rs6979218, which shows significant allelic activity only in proinflammatory, but not in resting, macrophages (**Figure 1h**). Together, our data indicates that both enhancer activity of elements as well as differential allelic activity of variants are influenced by biological contexts.

### Macrophage regulatory architecture undergoes rewiring in a proinflammatory context

Given the differences in the regulatory activity of AD GWAS variants upon proinflammatory stimulation, we hypothesized that deciphering the endogenous changes in transcriptional regulatory networks will be critical to understanding the regulatory impact of emVars. Therefore, we leveraged our previously published transcriptional regulatory networks across eight-point time courses of THP-1 macrophage activation^31^ (**Figure 2a**). We re-analyzed Hi-C, ATAC-seq, H3K27ac ChIP-seq, and RNA-seq datasets from resting (0h) and proinflammatory (24h) macrophages to identify changes in transcriptional regulatory networks that correspond to the timepoints used in our MPRA experiments. We performed differential analysis for each of these genomic datasets. In total, we identified 25,470 total chromatin loops with 1.61% (409) being differential (FDR < 0.05, log_2_(fold-change) [LFC] > 1), 163,150 ATAC-seq peaks with 12.5% (20,391) being differential (FDR < 0.05, LFC > 2), and 101,834 H3K27ac ChIP-seq peaks with 47.97% (48,850) being differential (FDR < 0.05, LFC > 2). Finally, we identified 20,738 genes, and 20.6% (4,273) of them were differentially expressed (FDR < 0.05, LFC > 2; **Figure 2a**). A subset of genomic loci display concordant increases in all of these genomic features during inflammation, deconvolving context-specific regulatory mechanisms. For example, at the *MARCKS* locus, increased contact frequency, chromatin accessibility, and histone H3K27ac is accompanied by elevated gene expression (**Figure 2b**).

**Fig. 2.**
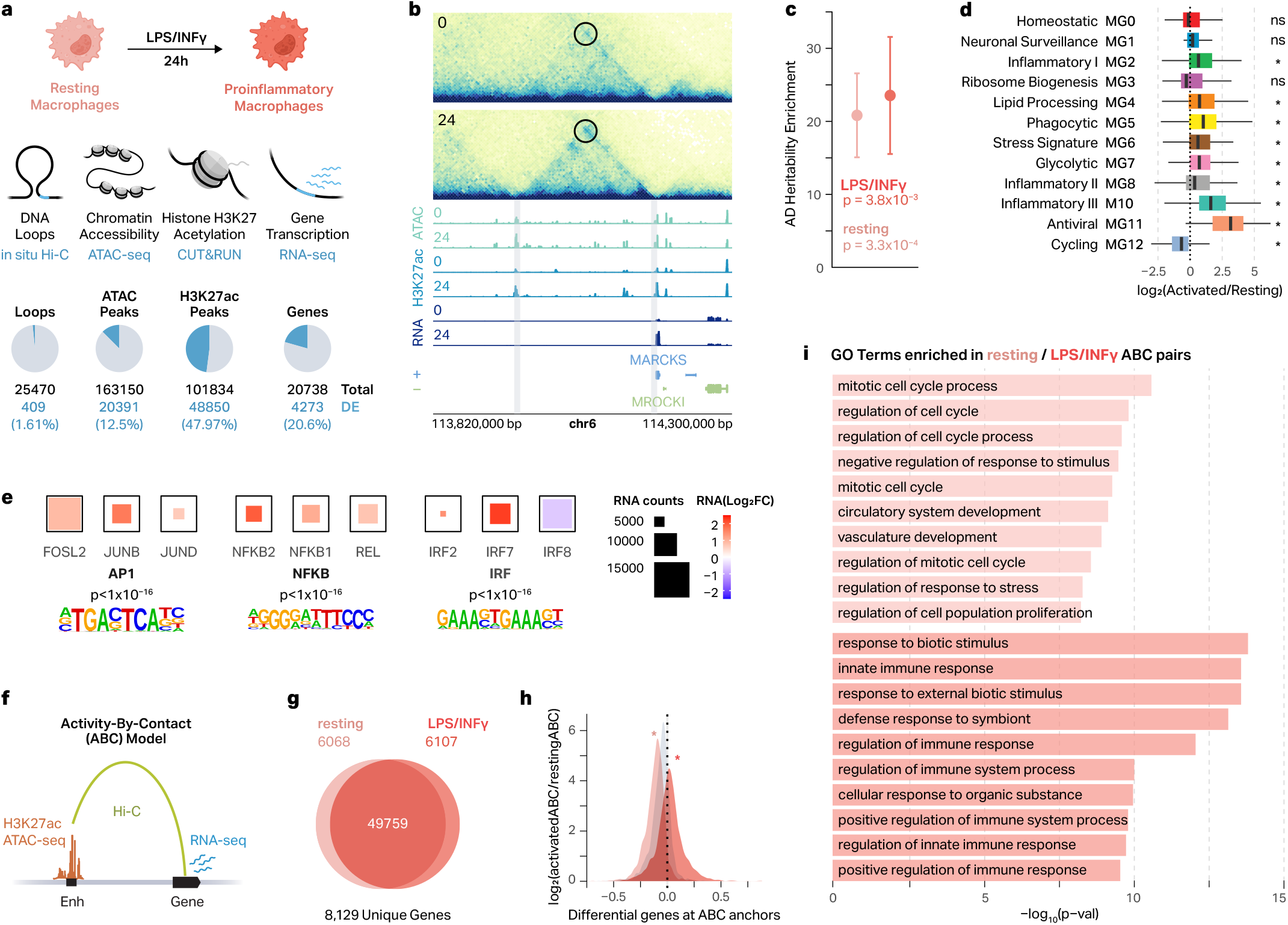
Macrophage regulatory architecture is rewired after proinflammatory response. **a**, Schematic of LPS+INFγ-treatment to generate proinflammatory macrophages and genomic data analyzed (top). Pie charts showing the number of differential loops, ATAC peaks, H3K27ac peaks, and genes between resting and LPS+INFγ-treated macrophages (bottom). Differential loops were considered significant with DESeq2 absolute LFC > 1 and FDR < 0.05. Differential genes and peaks were considered significant with DESeq2 absolute LFC > 2 and FDR < 0.05. **b**, Differential chromatin structure and gene expression at the *MARCKS* locus. 0, resting macrophages; 24, LPS+INFγ-treated macrophages for 24 hours. **c**, ATAC-seq peaks in resting and LPS+INFγ-treated macrophages are enriched for AD genetic risk factors. P-values calculated by s-LDSC. **d**, Box plots show the RNA LFC for LPS+INFγ-treated vs resting macrophages for genes in 12 scRNA-seq microglial clusters from AD postmortem brains. Box plots show the median and IQR with whiskers extending to the most extreme non-outliers. Asterisks (*) represent significance as calculated by a two-sided Wilcoxon rank-sum test (FDR < 0.05). **e**, TF motif enrichment for differential ATAC peaks in LPS+INFγ-treated macrophages and the expression of the top three most highly expressed members of those TF families. **f**, Schematic of the ABC model. **g**, Venn diagram showing the number of ABC-prioritized enhancer-gene pairs (ABC score > 0.05) identified in resting and LPS+INFγ-treated macrophages. **h**, Density plot showing the LFC in ABC score for genes highly expressed in resting macrophages (peach), genes highly expressed in LPS+INFγ-treated macrophages (salmon), or expression-matched static genes (gray). Asterisks represent significance as calculated by a two-sided Wilcoxon rank-sum test, proinflammatory vs static p = 1.3 x 10^−194^; resting vs static p = 2.2 x 10^−116^. **i**, GO terms enriched for DEGs at the anchors of resting- or LPS+INFγ-specific ABC pairs.

While it has been shown that regulatory elements of immune cells are enriched for genetic risk factors for AD^17,37^, the impact of regulatory element rewiring under proinflammatory conditions on this enrichment remains poorly understood. Therefore, we performed stratified linkage disequilibrium score (s-LDSC) regression analysis on ATAC-seq and H3K27ac ChIP-seq peaks from resting and proinflammatory macrophages. Both resting and proinflammatory ATAC-seq (and H3K27ac) peaks were enriched for AD-associated variants (p < 0.05, **Figure 2c, Sup Figure 3a**). As microglia are the major immune cell population in the brain, we compared the gene expression profiles of our resting and proinflammatory macrophages to the single-nucleus (sn)RNA-seq dataset of microglia from postmortem brains with and without AD^38^. Upregulated genes in proinflammatory macrophages were highly enriched for microglial clusters associated with inflammation (MG2, 8, 10), lipid processing (MG4), phagocytosis (MG5), stress (MG6), and antiviral (MG11) states (Two-sided Wilcoxon rank-sum test, FDR < 0.05). Conversely, upregulated genes in resting macrophages were enriched for a microglial cluster (MG12) associated with cell cycling (Two-sided Wilcoxon rank-sum test, FDR < 0.05, **Figure 2d**). Together, these results support the idea that transcriptional regulatory networks of proinflammatory macrophages could offer valuable insights into understanding molecular pathology of AD.

To further investigate the mechanisms underlying changes in differential chromatin architecture, we performed TF motif enrichment analyses. Regulatory regions more accessible in proinflammatory macrophages were strongly enriched for AP-1, NFKB, and IRF family members (**Sup Figure 3b, right**). Analysis with our matched RNA-seq data revealed that many of the members of these TF complexes were also upregulated in proinflammatory macrophages, including members of the AP-1 family (FOSL2, JUNB, JUND), NFKB complex (NFKB1, NFKB2, REL), and IRF family (IRF2, IRF7, IRF8; **Figure 2e**). Regulatory regions more accessible in resting macrophages were highly enriched for ETS, RUNX, and ELK family TF member motifs (**Sup Figure 3b, left**), which was not necessarily accompanied by decreases in the expression of the genes in those families (**Sup Figure 3c**). Importantly, many of the TF motifs enriched in differentially accessible chromatin regions were also enriched in condition-specific MPRA-active elements (**Figure 1d, Sup Fig 3b**), suggesting that differential chromatin architecture and differential MPRA activity may be regulated by the same mechanisms.

Using the Activity-By-Contact (ABC) model^39^ (**Figure 2f, Sup Table 5**), we identified 61,934 pairs amongst our resting and proinflammatory macrophages connecting to 8,192 unique genes. A majority of these pairs were shared between both conditions (49,759 enhancer-gene pairs, 80.3%), yet we still identified a substantial number of enhancer-gene pairs that were unique to each condition. For resting macrophages, we identified 6,068 (9.7%) unique enhancer-gene pairs, and for proinflammatory macrophages, we identified 6,107 (9.9%) unique enhancer-gene pairs (**Figure 2g**). Differential genes at proinflammatory-specific and resting-specific ABC anchors showed concordant upregulation and downregulation of gene expression respectively (Two-sided Wilcoxon rank-sum test, p < 0.05; **Figure 2h**).

We next asked whether the genes uniquely identified in resting- and proinflammatory-specific ABC pairs were relevant to macrophage immune responses. To address this, we performed gene ontology (GO) enrichment on these sets of genes. Genes at proinflammatory-specific ABC anchors were enriched for pathways involved in cytokine response and immune system regulation, consistent with the enrichment of NFKB and AP-1 motifs at accessible chromatin in the activated state (**Figure 2i**). Conversely, genes at resting-specific ABC anchors were enriched for pathways related to cell cycle regulation, consistent with the cells being in an unperturbed resting state prior to their transition into a proinflammatory phenotype (**Figure 2i**). Together, this suggests that the regulatory architecture of macrophages is rewired when macrophages become proinflammatory, shifting from genes involved in homeostasis to those involved in the inflammatory response.

### MPRA-activity correlates with cell type-specific chromatin accessibility and H3K27ac

We next assessed how MPRA-measured transcriptional activity compares with endogenous regulatory architecture in resting and proinflammatory macrophages. We divided our AD-associated genetic elements into 10 quantiles based on their MPRA-activity (**Sup Fig 4a**). We then determined the chromatin accessibility or H3K27ac dip score^40^ of each AD-associated genetic element to assess their endogenous enhancer activity. We found that AD-associated genetic elements with the strongest MPRA-measured transcriptional activity (elements in the 10th quantile) showed an increase in both accessibility and dip score in both resting and proinflammatory macrophages (Two-sided Wilcoxon rank-sum test, p < 0.05, **Figure 3a, Sup Fig 4b**). Furthermore, MPRA-active elements had higher dip scores and accessibility than MPRA-inactive elements in both conditions (**Sup Fig 4c-d**). This supports the notion that the strongest MPRA-active elements reside within accessible chromatin regions that likely function as enhancers. We were interested in understanding whether this trend was specific to macrophages, or was also apparent in other cell types. To address this, we obtained ATAC-seq data from the Roadmap Epigenomics study^41^ and extracted ATAC-seq counts for each AD-associated genetic element in various cell types. While elements in the 10th quantile showed enhanced chromatin accessibility across all cell types, macrophages exhibited the highest level of chromatin accessibility (**Figure 3b**).

**Fig. 3.**
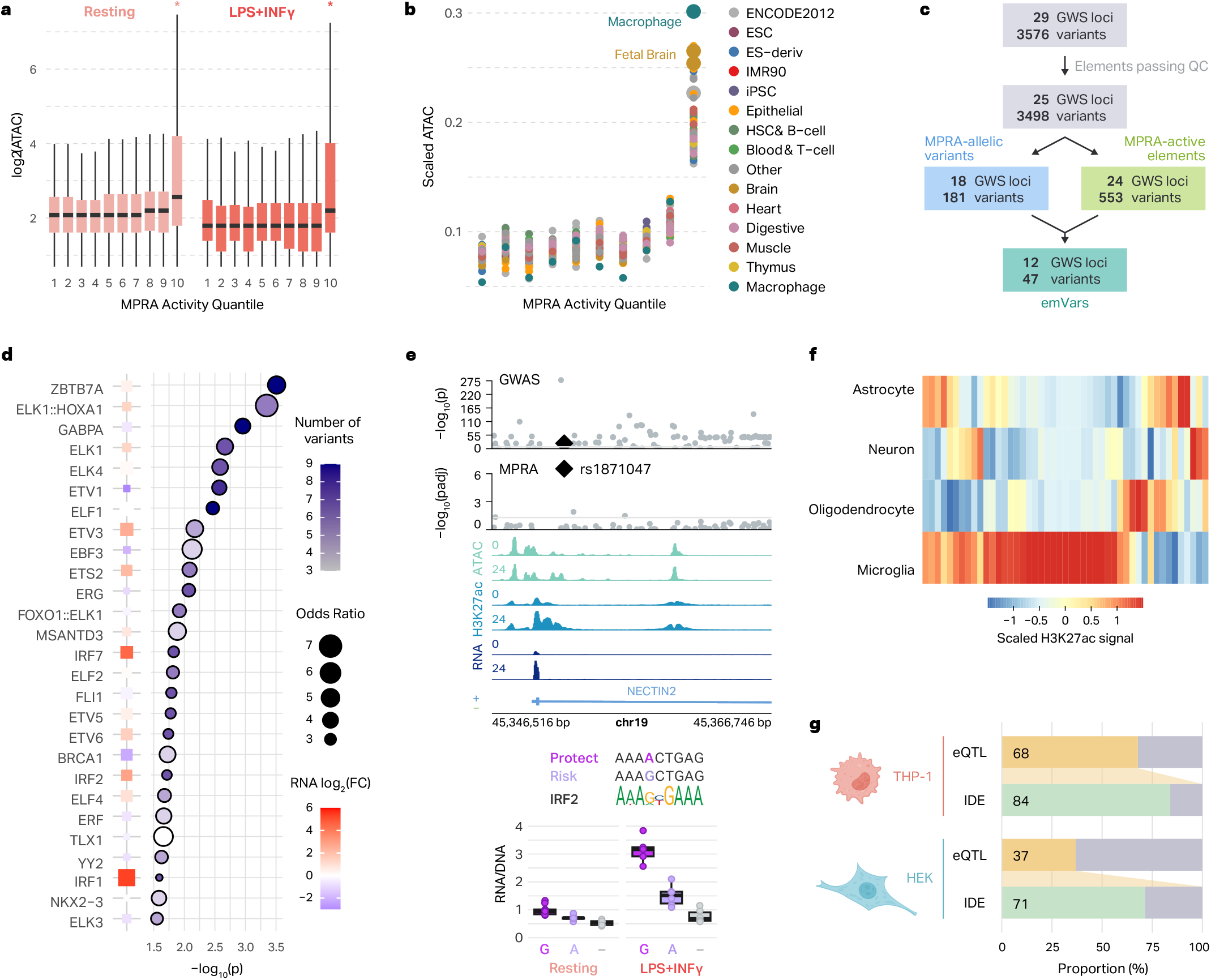
MPRA-measured regulatory activity correlates with endogenous regulatory architecture of macrophages and microglia. **a**, Box plots show the chromatin accessibility of AD-associated variants stratified by their MPRA-measured activity. Asterisks represent two-sided Wilcoxon ranksum test p < 0.05. The log_2_(ATAC-seq counts) for the variants in each MPRA activity quantile are displayed for the resting (left) and LPS+INFγ-treated (right) macrophages. Box plots show the median and IQR with whiskers extending to the most extreme non-outliers. **b**, Normalized ATAC-seq counts for AD-associated genetic elements are shown across MPRA activity quantiles for different cell types from the Roadmap Epigenomics study^37^. **c**, Workflow illustrating how emVars are defined. **d**, TFs with motifs predicted to be altered by emVars. Enrichment was calculated by comparing TF disruption in emVars with that in MPRA-inactive and MPRA-non allelic variants. The color of each point represents the number of emVars that disrupt the motif and the size of the point represents the odds ratio (right). P-values were calculated with a two-sided Fisher’s exact test. TF expression is compared between LPS+INFγ (red) and resting (blue) macrophages (left, as described in RNA LFC). **e**, Example region of an emVar that disrupts TF motif binding at the *NECTIN2* locus (top). Box plots show the transcriptional activity of each allele for the emVar for both resting and LPS+IN-Fγ-treated macrophages compared to negative control (–) (bottom). Box plots show the median and IQR with whiskers extending to the most extreme non-outliers. **f**, Heatmap of H3K27ac signals across four brain cell types at emVars. **g**, Comparison of microglia eQTL overlap with emVars defined in THP-1 macrophages and HEK293T cells.

This suggests that there is some cell type specificity that may contribute to the activity of an element tested in MPRA. Since Geschwind and colleagues previously performed MPRA on AD GWAS variants in HEK293T cells^42^, we compared our results to this independent dataset. Of the 3,494 variants that we assayed in THP-1 macrophages, 1,155 (33%) were also assayed in HEK293T cells (**Sup Fig 5a**). Of those, 73 (6.3%) and 107 (9.3%) were MPRA-allelic in THP-1 macrophages and HEK293T cells, respectively (**Sup Fig 5b**). Strikingly, only 7 variants showed allelic regulatory activity in both cell types, meaning that the majority of variants (90.4% for THP-1 macrophages and 93.5% for HEK293T cells) showed allelic activity in their respective cell types (**Sup Fig 5c**). Five out of the 7 variants had identical direction of effects (IDE) between the protective and risk alleles, and 2 had opposite directions of effect (**Sup Fig 5d**). In addition to allelic activity of variants, we compared MPRA-measured transcriptional activity of AD-associated genetic elements in both cell types. The most active AD-associated genetic elements in THP-1 macrophages were more accessible in THP-1 ATAC-seq data compared to the most active variants in the HEK293T MPRA (**Sup Fig 5e**). Together, this highlights the cell type specificity of AD-associated variants and the importance of conducting MPRA experiments in disease-relevant cell types.

### emVars disrupt TF motifs involved in homeostasis and inflammation

Next, we explored the potential mechanisms through which emVars might be influencing enhancer activity. We identified 47 emVars by intersecting MPRA-allelic variants with MPRA-active elements (**Figure 3c, Sup Fig 2c**). We reason that emVars are the primary candidates for disrupting enhancer activity in macrophages among the variants we have assayed, making them more likely to encode causal AD risk variants. Since variant regulatory activity is thought to be mediated by TF motif alterations^29,43,44^, we used motifBreakR^45^ to determine how emVars impact TF binding motifs (**Sup Table 6**). All emVars were predicted to significantly impact at least one TF motif. ETS and IRF family motifs amongst others were enriched in the set of altered motifs (two-sided Fisher’s exact test, **Figure 3d**). ETS and IRF TFs are involved in hematopoietic cell maintenance and inflammation respectively^46–48^ consistent with the notion that AD risk variants could heighten neuroinflammation.

A representative example of this is present at the *NECTIN2* locus. The emVar rs1871047 is located in a putative enhancer within an intron of *NECTIN2*, and appears to become more active in response to LPS+IFNγ treatment. This is consistent with expression of *NECTIN2* itself which is highly upregulated upon stimulation with LPS/IFNγ (**Figure 3e, top**). A closer examination of the variant reveals that it is located within an IRF2 binding site, where the risk G allele promotes IRF2 binding and is associated with increased expression of the reporter gene (**Figure 3e, bottom**). The *IRF2* gene itself is upregulated upon LPS+INFγ treatment and is associated with proinflammatory responses (**Figure 2e**). At this locus, IRF2 may bind to the risk G allele and promote expression of *NECTIN2*, an effect that is more pronounced upon an inflammatory response. *NECTIN2* encodes a lipid-related protein that is a component of adherens junctions. Interestingly, this protein has been shown to mediate viral entry into cells^49^ and has previously shown to be associated with AD^50^.

We next explored H3K27ac signatures of emVars across four major brain cell types^13^. We found that emVars were most enriched with H3K27ac signatures in microglia compared to other brain cell types, consistent with microglia being the resident macrophages of the brain (**Figure 3f**). As emVars were enriched with microglia regulatory elements, we intersected them with a well-powered microglia eQTL dataset^51^. Notably, 68% of emVars showed eQTL signatures in microglia, with 84% of them showing IDE (**Figure 3g**). These results suggest that MPRA-validated variant effects mimic the regulatory signatures of microglia. On the contrary, only 37% of emVars identified in HEK293T cells^42^ showed eQTL signatures in microglia, with 71% of them showing IDE, further highlighting that the transcriptional effects of emVars are cell type-specific.

### Mapping emVars to genes with ABC and eQTLs

To gain deeper insights into how emVars affect the transcriptomic landscape in AD, we identified putative target genes of our 47 emVars using two gene mapping approaches. First, based on our previous finding that the ABC model can effectively link emVars to the target genes^29^, we overlapped these variants with our macrophage ABC enhancer-gene pairs, resulting in 173 pairs that connect 23 emVars to 60 genes (**Figure 4a, Sup Table 7**). Second, we intersected emVars with microglia eQTL dataset^51^ (**Figure 3g**), resulting in 124 variant-eGene pairs connecting 32 emVars to 29 eGenes (**Figure 4a, Sup Table 7**). Together, we connected 39 of 47 MPRA emVars to 76 unique target genes (hereafter referred to as MPRA-AD risk genes; **Figure 4a-b**). Each MPRA-AD risk gene was connected to an average of 2.3 emVars, while each emVar was connected to an average of 4.8 target genes (**Figure 4c**). GO and KEGG pathway enrichment revealed that MPRA-AD risk genes are involved in the MHC class II protein complex, lipoprotein particles, endocytosis, and antigen processing (**Figure 4d, Sup Fig 6a**). emVars varied in their distance to the transcription start site of their mapped target genes, with a mean distance of 107 kb (**Sup Fig 6b**).

**Fig. 4.**
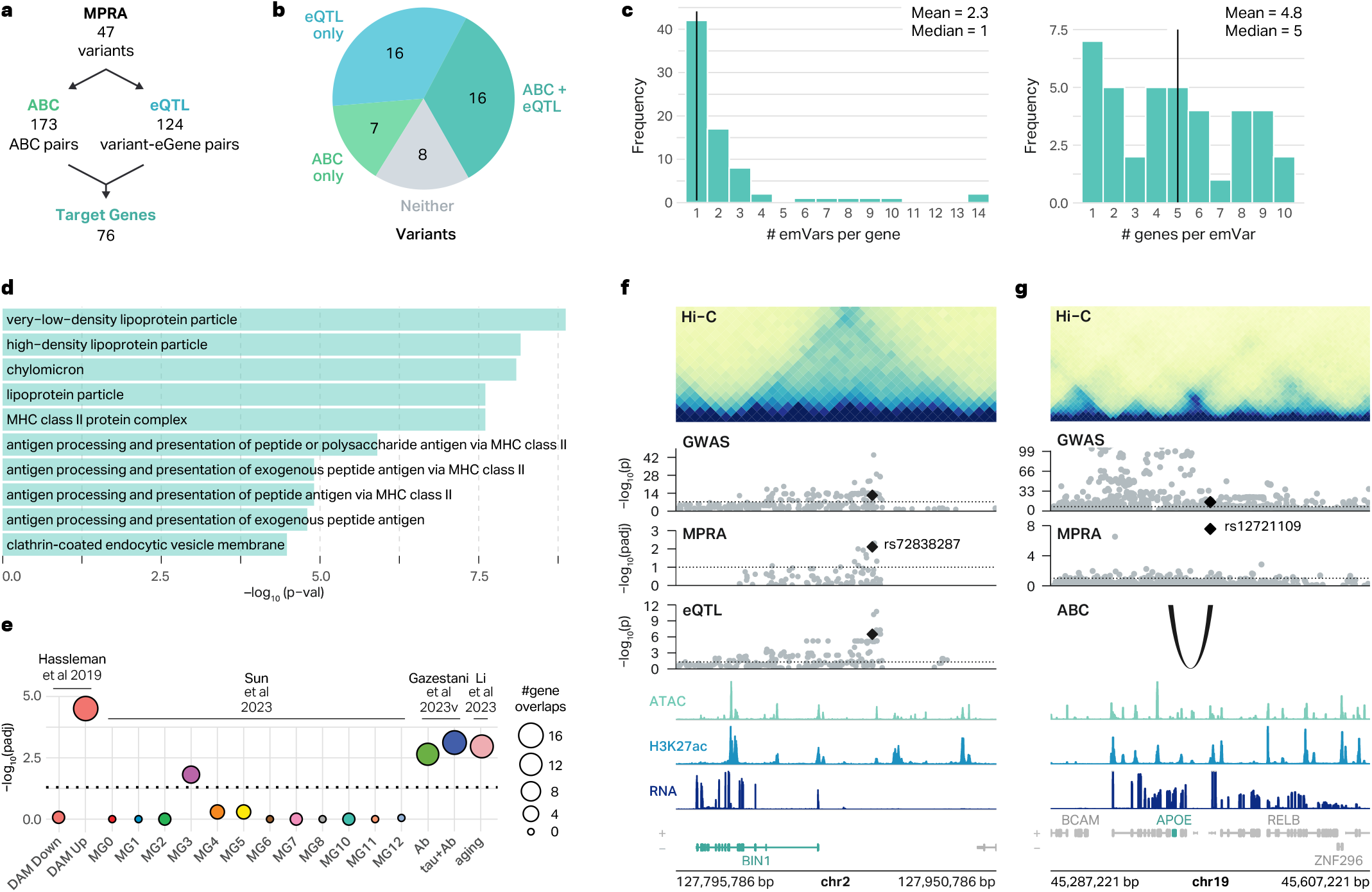
Mapping of emVars to AD risk genes. **a**, Workflow showing the number of emVars assigned to target genes by the ABC model and eQTLs. **b**, Pie chart showing the percentage of emVars that were assigned to a gene by the ABC model and eQTLs. **c**, Histogram showing the number of emVars connected to each MPRA-AD risk gene (left), and the number of genes connected to each emVar (right). **d**, GO terms enriched for MPRA-AD risk genes. **e**, Enrichment of MPRA-AD risk genes with (1) genes upregulated and downregulated in DAMs (Hassleman et al. 2019), (2) AD-associated microglia clusters (Sun et al. 2023), (3) genes differentially expressed in response to Aβ and Tau (Gazestani et al. 2023), and (4) microglial aging-related genes (Li et al. 2023). The size of the circle represents the number of MPRA-AD risk genes overlapping each gene signature. P-values were calculated with two-sided Fisher’s exact test. **f**, The emVar rs72838287 in the *BIN1* locus is connected to *BIN1* by microglia eQTLs. ATAC, H3K27ac, and RNA signal tracks have been merged across both conditions. **g**, The emVar rs12721109 in the *APOE* locus is connected to APOE by the ABC link in macrophages. ATAC, H3K27ac, and RNA signal tracks have been merged across both conditions.

We next explored the association of MPRA-AD risk genes with AD pathology by comparing these genes to various previously published datasets of AD-associated transcriptional landscapes. Given the emerging evidence implicating DAMs in AD pathology, we compared MPRA-AD risk genes with genes that are upregulated or downregulated in DAMs from the mouse brain^52^. We found significant enrichment of MPRA-AD risk genes among those that are upregulated, but not downregulated, in DAMs (two-sided Fisher’s exact test, FDR < 0.05, **Figure 4e**). We also leveraged 12 human microglial states defined from single-cell molecular atlas of the AD brains^38^ **(Figure 2d**). MPRA-AD risk genes were enriched for microglial state 3 (MG3), which involves ribosome biogenesis and exhibits the strongest DAM signature^38^ (two-sided Fisher’s exact test, FDR < 0.05, **Figure 4e**). In addition, MPRA-AD risk genes were enriched with differentially expressed genes in microglia in response to amyloid beta (Aβ) and tau^24^ (two-sided Fisher’s exact test, FDR < 0.05, **Figure 4e**). Finally, since AD reflects age-associated impairment, we evaluated whether MPRA-AD risk genes exhibit aging-dependent molecular signatures^53^. MPRA-AD risk genes were enriched with genes with aging-dependent molecular trajectories in mice between the age of 2 and 24 months (two-sided Fisher’s exact test, FDR < 0.05, **Figure 4e**). Importantly, 19 out of 76 MPRA-AD risk genes (25%) drove the enrichment of AD-associated gene signatures connected to 26 emVars, suggesting that genetic risk factors for AD may contribute to the molecular pathology present in AD brains.

MPRA-AD risk genes included well known AD risk genes such as *BIN1* and *APOE*. In addition to identifying putative target genes, our approach allowed us to prioritize variants and regulatory mechanisms that are likely driving the association of *BIN1* and *APOE* with AD. In the *BIN1* locus, we detected an emVar rs72838287 located in a putative macrophage enhancer marked by ATAC-seq and H3K27ac peaks (**Figure 4f**). Microglia eQTLs also supported the relationship between rs72838287 and *BIN1* expression. In both MPRA and eQTLs, risk allele G was associated with increased expression of GFP and *BIN1* respectively. *BIN1* has been previously reported to be involved in Tau-mediated neurotoxicity^54^, proinflammatory responses^55^, and neuronal activity and communication^56–58^. In the *APOE* locus, we found an emVar rs12721109 located within a putative macrophage enhancer supported by ATAC-seq and H3K27ac peaks (**Figure 4g**). Rs12721109 is connected to the promoter of *APOE* by a chromatin loop and a strong ABC connection. The risk allele G was associated with increased expression of GFP in our MPRA, suggesting that *APOE* may be upregulated in AD. In addition to the well-established role of *APOE* ε4 allele in increasing AD risk^59,60^, our results suggest that non-coding variants in the locus may also affect AD susceptibility by modulating *APOE* expression. In line with this hypothesis, *APOE* was found to be upregulated in response to Aβ and Tau^24^ and in DAMs^52^.

### An emVar shows allele-specific disruption of TF binding

It is thought that the variant regulatory activity measured by MPRA is largely mediated by TF motif disruption^29,44^. To test this, we performed EMSAs on rs9270887, an emVar at the HLA locus. This variant showed the largest difference in allelic activity between risk and protective alleles among all emVars identified at the HLA locus (**Figure 5a**). Both the ABC model and eQTLs suggest that the target gene of rs9270887 is *HLA-DRB1*, potentially providing a mechanism for the highly complex MHC locus associated with AD. The risk allele A was associated with increased sGFP expression in both resting and proinflammatory macrophages (**Figure 5b**).

**Fig. 5.**
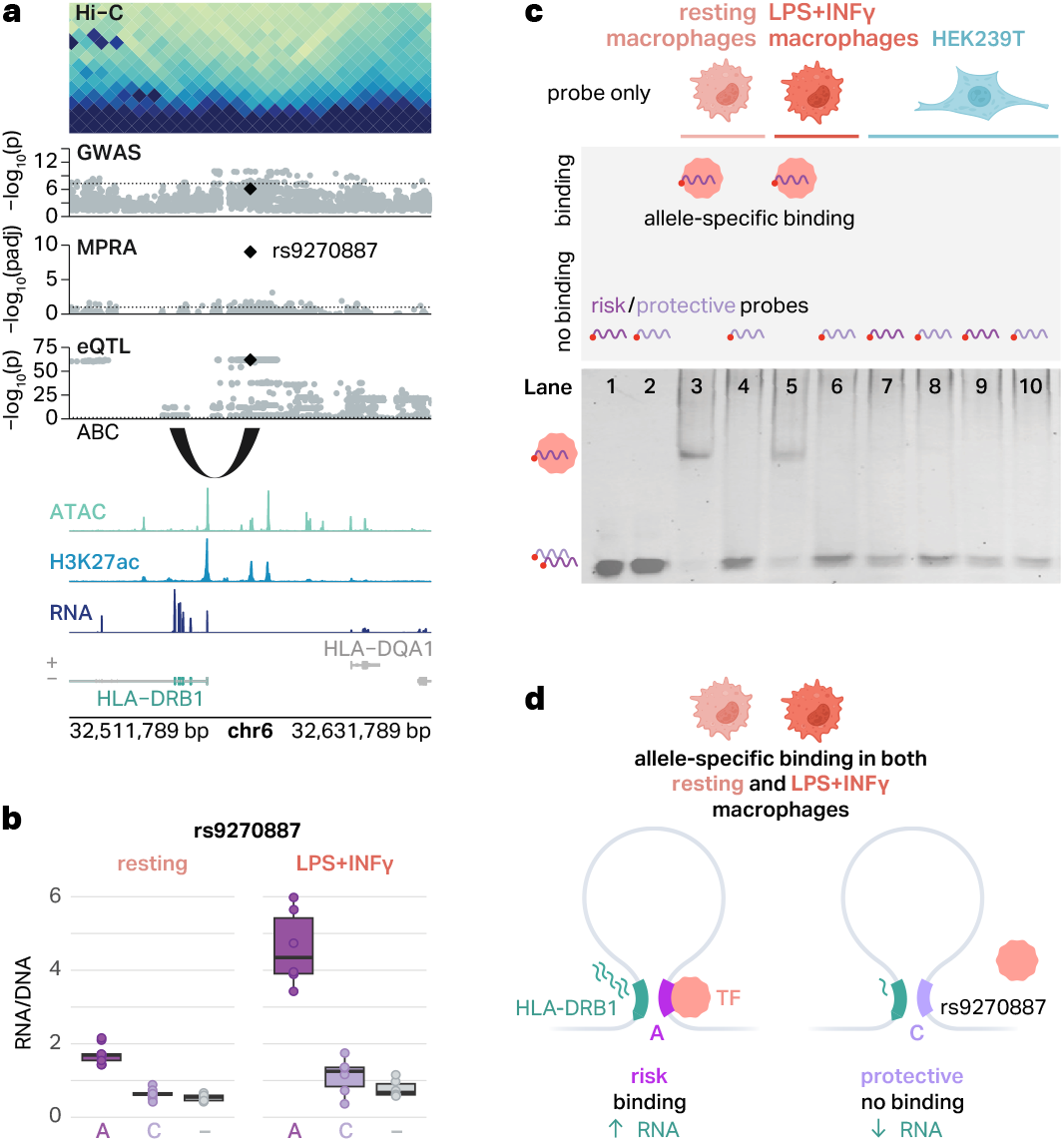
An emVar at the *HLA* locus shows cell type-specific and allele-specific protein binding. **a**, The emVar rs9270887 in the *HLA* gene cluster is connected to *HLA-DRB1* via the ABC model and eQTLs. ATAC, H3K27ac, and RNA signal tracks have been merged across both conditions. **b**, Box plots show the transcriptional activity of risk and protective alleles of rs9270887 in resting and LPS+INFγ-treated macrophages compared to negative controls (–). The boxplots show the median and IQR with whiskers extending to the most extreme non-outliers. **c**, Graphical representation of experimental conditions and bands detected in EMSAs (top) and EMSA results for the emVar rs9270887 (bottom). Lanes 1-2, risk (1) and protective (2) probes without lysates. Lanes 3-4, risk (3) and protective (4) probes with lysates from resting macrophages. Lanes 5-6, risk (5) and protective (6) probes with lysates from LPS+INFγ-treated macrophages. Lanes 7-8, risk (7) and protective (8) probes with HEK293T lysates. **d**, Schematic of our suggested mechanism of regulation of *HLA-DRB1*. In THP-1 macrophages, there is a chromatin loop connecting rs9270887 to the promoter of *HLA-DRB1*. The risk A allele leads to increased transcription of GFP, likely due to TF binding. The protective C allele breaks this TF binding site, which is associated with decreased transcription of GFP.

We designed 41 bp fluorescent probes encoding risk and protective alleles of rs9270887 and performed EMSAs using lysates from both resting and proinflammatory macrophages. We observed a shift in the band only for the risk allele in both resting and proinflammatory macrophages, suggesting that only the risk allele has a potential for TF binding (**Figure 5c**). This result is consistent with the MPRA finding where the risk allele, which preferentially binds to TFs in EMSAs, correlates with increased gene expression (**Figure 5b**). Notably, rs9270887 was previously tested in HEK293T cells but did not display significant allelic regulatory activity (p = 0.5)^42^. Therefore, we interrogated cell type-specific TF binding to this variant by performing EMSAs with HEK293T lysates. We found that neither the risk allele nor the protective allele binds to TFs in HEK293T lysates (**Figure 5c**), again supporting cell type-specific regulatory activity. This provides a compelling regulatory mechanism explaining how the risk allele for rs9270887 displays transcriptional activity in a cell type-specific fashion.

## Discussion

GWAS have identified loci associated with AD risk^3^, but interpretation is challenging because each locus contains dozens of closely linked risk variants due to LD. Furthermore, most variants are in non-coding regions, suggesting they have regulatory function. Therefore, recent studies attempted to identify causal AD variants and distill their functional impacts by intersecting AD GWAS variants with multi-omic datasets collected from primary microglia from AD brains^51^ as well as human induced pluripotent stem cell (hiPSC)-derived microglia^61^. While overlap between risk variants and functional annotations (e.g. chromatin accessible region, histone H3K27ac peak, etc) provides valuable insights into putative regulatory mechanisms, it does not provide direct evidence that a variant alters regulatory activity. MPRA has been used to directly quantify the regulatory impacts of AD-associated variants in HEK293T cells^42^ However, given that AD genetic risk factors show immune cell-specific heritability enrichment^13,18,16,17^ and that MPRA demonstrates cell type specificity^29,62–64^, it remains unclear whether the variant effects observed in HEK293T cells translate to AD-relevant biological contexts.

To address these limitations, we used MPRA to systematically characterize regulatory function of AD GWAS variants in THP-1 macrophages in both resting and proinflammatory states. To the best of our knowledge, this is the first study to perform MPRA on AD risk variants in a cellular model and biological context relevant to AD. We identified 47 AD risk variants that were significantly more active than negative controls and exhibited allele-specific differences in activity (emVars). We then reanalyzed our previously published RNA-seq, ATAC-seq, H3K27ac CHIP-seq, Hi-C, and eQTL data^31,51^ to identify 76 putative AD risk genes. These genes were enriched for genes previously implicated in AD.

Comparison of our results with those obtained from HEK293T cells^42^ highlighted the cell type specific nature of MPRA. Only 5 out of 1,155 variants tested in both studies were found to have shared allelic effects in both cell types. Moreover, ∼57% of our emVars showed eQTL signatures with IDE in microglia^51^, compared to ∼26% of emVars identified in HEK293T cells. These results collectively suggest that regulatory effects of emVars identified in THP-1 macrophages mimic regulatory signatures of microglia and may provide a more accurate quantification of the transcriptional impacts of AD risk variants. However, it is important to note that this difference could be attributed to different designs used in two studies. MPRA libraries in HEK293T cells were introduced episomally, while in our study, MPRA libraries were delivered to THP-1 macrophages via lentiviral infection, which results in integration and chromatinization within the host genome. It has been shown that the MPRA activities of chromosomally integrated sequences can differ from those of identical sequences assayed in episomes, with integrated sequences typically showing a better overlap with ENCODE enhancer annotations^65^. Therefore, further investigation is needed to determine whether this difference is attributable to differences in cell types or delivery methods.

Comparing our results to previous genomics-based studies allowed us to revise attributions of putative causal variants at several loci including the well studied AD risk gene *BIN1*. Previous studies have attributed AD risk causality to rs6733839^13^ and rs13025717^42^, and CRISPR-mediated perturbation of regulatory elements containing them did impact *BIN1* expression. However, in our MPRA analysis, these risk variants were not identified as emVars. The discrepancy in rs6733839 may stem from differences between allelic and enhancer activity because while the variant’s enhancer activity was tested using CRISPR-mediated perturbation, its allelic activity has not been previously assessed. The discrepancy for rs13025717 may be attributed to cell type specificity, as it exhibited enhancer activity but not allelic activity in THP-1 macrophages. We identified a novel emVar, rs72838287, with a regulatory connection with *BIN1* in THP-1 macrophages. These differences highlight the importance of testing enhancer and allelic effects of the variant in a relevant biological context.

In addition to cell type specificity, we examined variant regulatory activity under proinflammatory conditions to better model neuroinflammation in AD, as variants’ functional consequences can vary with different conditions^12–15^. Notably, ∼38% of MPRA-active elements and 68% of MPRA-allelic variants exhibited condition specificity, suggesting that variants may have condition-specific regulatory effects due to both alterations in the activity of the regulatory elements and changes in the direction of allelic activity.

These findings represent an important advance in understanding the genetic basis of AD, as we have identified at least one emVar in 12 of the 29 GWAS loci analyzed. However, it is important to recognize several limitations. First, while we interrogated the majority of AD GWAS risk variants known at the time, this study did not quantify the regulatory impact of all currently available AD risk variants. For technical reasons, we excluded variants that resulted in insertion or deletion as well as those that were not compatible with our cloning experiments. Moreover, since the inception of this project a larger AD GWAS study has been published that has increased the number of risk loci. Second, while many of the variants are thought to influence macrophages and microglia function, it is possible that some variants impact other cell types and/or biological states not tested here. Finally, while the variants and genes identified here have a high likelihood of influencing AD risk, proving and understanding their role in AD initiation and progression will require rigorous functional studies in animals and/or organoid models.

In summary, our work breaks down critical barriers to understanding and treating AD by quantifying the regulatory impacts of AD risk variants and revealing those that alter activity in both resting and proinflammatory macrophages. The next step will be to determine how these variants influence AD relevant phenotypes in multicellular/animal models while also characterizing variants identified in newer GWAS studies. Together, these steps will pave the way for novel therapeutic strategies and personalized interventions in AD treatment.

## Methods

### CELL CULTURE

#### Macrophage differentiation and activation

The THP-1 human monocyte cell line (ATCC, #ATCC® TIB-202TM), was cultured in RPMI 1640 media (Sigma-Aldrich # R8758) with the addition of 10% FBS (Thermo Fisher Scientific, Gibco TM, #10500064) and 1% penicillin/streptomycin (Sigma-Aldrich, #P4333). In order to differentiate THP-1 monocytes into macrophages, we plated them at a density of 1.0 × 10^6^ cells/well in 6-well plates and incubated with 25 μM PMA (12-O-tetradecanoylphorbol) for 24h. Macrophages were then allowed to rest for 72h before LPS/INFγ-treatment. A proinflammatory state was induced by treating the cells with 10 ng/mL of LPS (Sigma-Aldrich # L2630) and 20 ng/mL INFγ (Peprotech, #300-02) during 24h.

### MPRA

#### Variant selection and creation of oligo library

We have selected variants with nominal significance (p < 1×10^−5^) from 29 AD-associated GWAS loci^30^. Variants containing specific restriction enzyme sites for cloning purposes (MluI, SpeI, KpnI, XbaI) were excluded. Each variant was positioned at the center of a 150bp segment of its genomic context. Since GWAS variants were based on hg19, we used hg19 to retrieve the sequence. We included tiled CMV and EF1 promoters as positive controls and 198 randomly scrambled DNA sequences with matching GC content to the MPRA library as negative controls. Agilent oligoarray chip technology was utilized to synthesize 150bp sequences encompassing the variants along with flanking restriction enzyme sites for cloning, resulting in a total sequence size of 200bp.

#### Engineering of lenti-MPRA backbone

Plasmid pLS-mP (addgene Plasmid #81225) was modified to be used for the MPRA experiment. Since the pLS-mP plasmid contained SpeI and MluI restriction sites that would disrupt the downstream cloning of the MPRA library, we first removed these sites by digesting the plasmid with SpeI-HF (NEB, cat#R3133S), MluI-HF (NEB, #R3198S), and rSAP (NEB, #M0371S), followed by ligation with Remove-spel-mlul oligo (**Supplementary Table 1**) using Quick Ligase (NEB, #M2200S). The modified plasmid was transformed into electrocompetent cells (Endura, cat#60242), and plated on lysogeny broth (LB) agar plates with ampicillin (ThermoFisher Scientific, #J60977.14). Several colonies were selected and mini prepped using QIAprep Spin Miniprep Kit (Qiagen, #27104). These colonies were digested with SpeI-HF and XbaI (NEB, #R0145S) to check for the correct insert of the Remove-spel-mlul-R oligo. Colonies with the correct insert were used for downstream modification.

Next, a KpnI restriction enzyme site needed to be removed, so it would not disrupt the downstream cloning of the MPRA library. The plasmid was first digested with Bsu36I (NEB, #R0524S), KpnI-HF (NEB, #R3142S), and rSAP. Remove-KpnI-oligo (**Supplementary Table 1**) was ligated into the digested plasmid using Quick Ligase. The plasmid was transformed into electrically competent cells and selected using the aforementioned method. The KpnI restriction site removal was verified by digestion of the plasmid with KpnI-HF and XbaI digestion. Colonies with the correct insertion were used for downstream modification.

Finally, the plasmid’s minimal promoter and eGFP were removed and new SpeI and MluI restriction sites were added in. The plasmid was digested with EcoRI-HF (NEB, #R3101S), XbaI, and rSAP. The correct sized band was gel extracted using Zymoclean Gel DNA Recovery Kits (Zymo, #D4008). The Add-Spel-Mlul oligo (**Supple mentary Table 1**) was ligated into this site using Quick Ligase. The plasmid was transformed into electrically competent cells and selected using the aforementioned method. We validated the correct insert by digesting with BspEI (Kpn2I) (NEB, #R0540S) and MluI, and with sanger sequencing of the region. Colonies with the correct insert served as the plasmid backbone for the MPRA library generation.

#### Inserting variant library into lenti-MPRA backbone

The lyophilized 200bp sequences from Agilent were resuspended in TE buffer at 2 pM, and PCR primers (MPRA-chiprimer-F3 and MPRA-chiprimer-R3, sequences available in **Supplementary Table 1**) were used to amplify the AD variant library and controls. PCR reactions were cleaned using Zymo PCR clean and concentrator-5 (Zymo, #D4003). Biotinylated PCR primers (MPRA-bcprimer-F, MPRA-bcprimer-R, sequences available in **Supplementary Table 1**) were used to add random 20bp barcodes to each sequence as well as SpeI and XbaI restriction enzyme sites for downstream cloning. The library was subsequently digested with SpeI-HF (NEB_#R3133L) and MluI-HF (NEB_#R3198S) for 1h at 37°C and bead cleaned with Dynabeads MyOne Streptavadin C1 beads (Thermo Fisher Scientific_#65001) to recover non-biotinylated library-barcode pairs. We also digested 5ug of the modified pLS-mP backbone plasmid with SpeI-HF, MluI-HF (NEB_#R3198S), and rSAP for 3h at 37°C, which is followed by heat inactivation for 20 minutes at 80°C, and gel extraction using Zymo Gel DNA Recovery kit (Zymo_#D4008). The digested pLS-mP backbone and library-barcode pairs were ligated using T7 DNA ligase (NEB_#M0318S) at room temperature for 30 minutes. The ligated product was immediately transformed into electrocompetent cells and grown at 30°C overnight on LB agar plates with ampicillin. Colonies were quantified to estimate barcode complexity, and all colonies were scraped and grown in LB with ampicillin at 30°C overnight. The culture was maxi prepped using Qiagen’s Plasmid Maxi Kit (Qiagen 12162). To check that the library was inserted, we performed (1) colony PCR (primers: P7_Lenti_F and P5_Lenti_R, sequences available in **Supplementary Table 1**) and (2) restriction enzyme digest using XbaI and PstI-HF. This resulted in the AD-MPRA-variant-barcode library.

#### Barcode mapping

The variant and barcode region of the AD-MPRA-variant-barcode library was amplified via PCR using the NEBNext 2X Q5 Hifi HS Mastermix (NEB, #M0541L) and primers containing Illumina P5 and P7 adapters (Bcmap_P5_R and Bcmap_P7_F, sequences available in **Supplementary Table 1**). The PCR product was cleaned up using the Zymo PCR clean and concentrator-5 kit. The resulting library was sequenced using custom sequencing primers (BCmap_R1Seq_R and BCmap_R2Seq_F, sequences available in **Supplementary Table 1**) with the paired-end 250bp Novaseq 6000 SP platform at the UNC High-Throughput Sequencing Facility (HTSF). Barcodes were assigned to each variant using a custom code currently available in a github repository (https://github.com/kiminsigne-ucla/bc_map) as previously described^29^.

#### Adding in minimal promoter and GFP

We amplified a minimal promoter (minP) and GFP (minP-GFP) from the unmodified original pLS-mP plasmid (https://www.addgene.org/81225/) via PCR (primers: minP-GFP-F and minP-GFP-R, sequences available in **Supplementary Table 1**) using the NEBNext 2X Q5 Hifi HS Mastermix. The amplified minP-GFP fragment was then cleaned up using the Zymo PCR clean and concentrator-5 kit. The minP-GFP fragment and variant-barcode library were both digested with KpnI-HF (NEB, #R3142S) and rSAP for 3h at 37°C, which was followed by heat inactivation for 10 minutes at 65°C. Both of these products were then gel extracted from a 0.8% agarose gel using Zymo Gel DNA Recovery kit. The gel extracted products were then digested with XbaI (NEB, #R0145S) and rSAP for 3h at 37°C, and heat inactivated for 10 minutes at 65°C. Digested minP-GFP and variant-barcode library plasmid were cleaned up using Zymo PCR clean and concentrator-5 and ligated together using T7 DNA ligase. The ligation mix was, incubated at room temperature for 30 minutes, then cleaned up using Zymo PCR clean and concentrator-5. The ligation mix was transformed into electrocompetent cells, which were then plated on LB agar plates with ampicillin, resulting in the AD-MPRA library. The AD-MPRA library was grown in LB with ampicillin, and maxi prepped using Qiagen Maxi prep kit. The UNC Gene Therapy Center packaged the AD-MPRA library into lentivirus in two batches. The resulting virus had the titer of 4.56×10^9^ viral particles/µL and 2.05×10^10^ viral particles/ µL respectively.

#### Lentiviral transduction of THP-1 cells

THP-1 cells were transduced with AD-MPRA libraries five days prior to the start of the differentiation into macrophages. Briefly, THP-1 cells were counted and resuspended at a density of 1 million cells per mL in fresh media with FBS and 8 μg/mL of polybrene. 110-300 lentiviral particles per cell were used for transduction. This number was determined based on titration experiments that assessed cell viability and transduction efficiency for each independent lentivirus batch. Through these titration experiments, we selected the number of viral particles per cell that achieved the maximum transduction efficiency without significantly compromising cell viability. After the addition of the lentivirus, cells were centrifuged for 30 minutes at 800g at 32°C and plated on 12-well plates. A no-virus control was included to monitor general cell health. After 20h, cells were centrifuged for 5 minutes at 300g, old media with lentivirus was removed, and cells were resuspended in fresh media and plated on 12 well/plates at a density of 4×10^5^ cells/mL. Cells were left to recover for 4 days and on day 5, macrophage differentiation was initiated. RNA was extracted from each well 10 days after transduction. To enhance detectability of transduced cells, we pooled 1.2-1.8 million cells per replicate.

#### DNA and RNA isolation and processing

RNA was extracted using the RNAeasy mini kit (Qiagen, #74104) following manufacturer’s instructions. RNA integrity number (RIN) was measured for all samples using Agilent tapestation 4150 system. RNA samples with a RIN score ≥ 8.5 were employed. Barcode cDNA was generated from the total extracted RNA using SuperScript IV Reverse Transcriptase (Invitrogen, #18090050) and a primer that targets downstream of the barcodes (Lib_Hand_RT_ Lenti, sequences available in Supplementary Table 1). To acquire an initial input of DNA barcodes introduced into the cells, RNA was extracted from the cell-free lentivirus preparation which contained the AD-MPRA library using the NucleoSpin Virus kit (Takara, #740983.10). The viral RNA was retro-transcribed using SuperScript IV Reverse Transcriptase and the Lib_Hand_RT_Lenti primer. This DNA was used to quantify MPRA DNA barcodes for normalization.

#### RNA and DNA MPRA library preparation

cDNA from the cell-free AD-MPRA library (lentivirus) and cDNA from each transduced well were amplified via PCR using the Q5 High fidelity 2X Master Mix (NEB, #M0492S) and Lib_Hand_primer and Lib_Seq_GFP_R_ primers (sequences available in Supplementary Table 1). The samples were cleaned up using the DNA clean and concentrator-5 kit. Next, a second amplification step was performed to add on sequencing adaptors and unique Illumina indices (#). For this, the Q5 High fidelity 2X Master Mix was used again with the P5_Seq_GFP_Lenti_F and P7_Ind_#_Han primers (sequences available in Supplementary Table 1). The resulting libraries were cleaned up using 1:1 ratio of Ampure XP beads (Beckman Coulter, #A638819) and sequenced by the UNC HTSF using the NovaSeq 6000 S1 platform and at New York Genome Center using the NovaSeq X 10B platform (1×35bp) with custom primers that capture the barcode sequence and sequencing index (read 1 primer: Exp_R1_seq_P, index primer: Exp_Ind_seq_P).

#### Electrophoretic Mobility Shift Assay (EMSA)

HEK293T cells were cultured in DMEM containing GlutaMAX (Thermo Fisher Scientific, #10566016), 10% FBS (Fisher Scientific, #26-140-079), and 1% sodium pyruvate (Thermo Fisher Scientific, #11360070). THP-1 cells were differentiated into macrophages and stimulated with LPS and IFNγ as described above. Nuclear lysates from control and stimulated macrophages, and HEK293T cells were extracted using the NE-PERTM Nuclear and Cytoplasmic Extraction Kit (Thermo Fisher Scientific).

5’ IRDye 700 labeled duplexed oligonucleotide probes (rs9270887_risk and rs9270887_protect, sequences available in **Supplementary Table 1**) with centrally positioned variants were obtained from Integrated DNA Technologies. EMSAs were carried out with the Odyssey Infrared EMSA kit (LICORbio, #829-07910) according to the manufacturer’s instructions. Each binding reaction consisted of 10 µg of nuclear extract combined with 2 µl of 10X binding buffer, 1 µl of 1 ug/µl poly(dl-dC), 2 µl of 25 mM DTT/2.5% Tween 20, 1 µl of 1% NP-40, 1 µl of 50 nM IRDye 700-labeled probe and water to a total volume of 20 µl. The reactions were incubated at room temperature protected from light for 1h. To confirm specificity of the band resulting from protein-probe complex, unlabeled duplex DNA (45X) was added for competition reactions. Prior to loading the samples, 5% TBE gel (Bio-Rad) was pre-run in the 0.5X TBE running buffer for 45 minutes. Following electrophoresis, DNA-protein complexes were detected using the Azure 600 imaging system.

## Data Analysis

### MPRA DATA ANALYSIS

#### MPRA data processing

Using the barcode-variant definitions from barcode mapping, RNA barcodes from each replicate in each condition were mapped to their corresponding variants and were counted. Since one DNA sample was used for all replicates in each condition, barcodes were connected to their corresponding variants for the DNA as well. Variants were found to have mapped to a median of 107 barcodes. A count matrix was constructed containing the RNA barcode counts across all samples with the DNA barcode counts. Barcodes that were defined as outliers were removed, and variants with fewer than 5 barcodes mapped were removed as well. For each variant, the RNA and DNA barcode counts were aggregated. We conducted reproducibility analysis between each of the 9 resting biological replicates and 6 proinflammatory biological replicates.

#### Identification of active elements

The aggregated RNA and DNA counts were used to calculate a RNA/DNA ratio to quantify MPRA element activity. The scrambled negative controls were further aggregated to result in one single negative control RNA/DNA ratio per each of the 9 resting biological replicates and each of the 6 resting biological replicates for a total of 15 biological replicates. A fixed linear mixed-effect model (lmer function from the lme4 R package, version 1.1.35.1) was used to determine which AD-associated genetic elements had significantly higher activity than negative controls. The following design was used: activity ∼ var*condition + (1 | pairs). In this design, “var” is whether that element is an MPRA-tested element or a negative control, “condition” is the biological context (resting or LPS+INFγ), and “pairs” is the replicate being tested (this is to account for differences in sequencing depth across replicates).

To identify MPRA-active elements, we selected the allele with the smallest p-value for the “var” coefficient and applied a Benjamini-Hodgeberg p-value correction. MPRA-active elements were defined as those with an FDR < 0.05 and a beta > 0 (meaning that the element is more active than the negative controls).

To define MPRA-active elements specific to one context, we first selected MPRA-active elements and applied a Benjamini-Hodgeberg correction to the p-value from the interaction of var*condition in the above model. Resting-specific elements had an FDR < 0.05 and an interaction beta < 0; LPS+INFγ-specific elements had an FDR <0.05 and an interaction beta > 0.

#### Identification of allelic regulatory variants

Using the aggregated RNA and DNA counts from above, we used the mpralm function from the mpra^66^ (version 1.20.0) R package to calculate differential allelic regulatory activity. The following parameters were used: mpra_lm_object <-mpralm(object = mpra_set, design = design_matrix, aggregate = “none”, normalize = T, block = samples, model_type = “corr_groups”). The mpra_set object is an mpra object that was generated with the MPRAset function, which uses DNA and RNA barcode counts per allele as the input. To maximize power, the biological replicates for both conditions were combined together, resulting in 15 biological replicates with the condition effect regressed out in our design_matrix. The resulting logFC was adjusted to account for GWAS effect size (called “corrected_logFC” in supplemental tables), resulting in variants with a logFC > 0 being associated with AD risk, and variants with a logFC < 0 being associated with AD protection. MPRA-allelic variants were defined as those that have a FDR< 0.05 for the difference in activity between the risk allele and protective allele.

To identify variants with an interaction between condition and allele, we subsetted the mpralm result object for the interaction term coefficient and subsetted for all 181 MPRA-allelic variants. We applied a Benjamini-Hodgeberg correction and designated variants with FDR < 0.05 as significant.

#### Expression-modulating variant (emVar) identification

We defined expression-modulating variants (emVar) as all MPRA variants that were MPRA-active for at least one allele in at least one condition and identified as an MPRA-allelic variant.

#### TF motif enrichment analysis

TF motif enrichment was performed on the 150 bp sequences surrounding resting- or proinflammatory-specific MPRA elements using the findMotifsGenome.pl function from HOMER (version 4.11). Sequences that did not change in activity between contexts were used as background peaks for both sets. The default parameters were used with “-size given”.

### GENOMIC DATA ANALYSIS

#### Hi-C Data processing and differential loop calling

Hi-C data was processed using a modified version of the Juicer pipeline^10^ (version 1.5.6) and aligned to the hg19 genome build. Hi-C maps were generated at 5, 10, 25, 50, 100, 200, 250, 500, 1000, and 2500 kb resolution for 3 biological replicates and 2 conditions for each replicate. Replicate 1 had 1 technical replicate, replicate 2 had 2 technical replicates, and replicate 3 had 2 technical replicates. This resulted in 10 unique samples total. Each of the 5 hic files from each condition were combined into one Hi-C map per condition. Finally, all of the 10 samples were combined to create one “Mega” map as described in Reed et al 2022^31^.

Loops were called from the merged condition and Mega Hi-C map with SIP^67^ (version 1.6.2) with settings “-g 2 -5 2000 -fdr 0.05” at 5 kb resolution with KR normalization factors. A count matrix was generated by extracting unnormalized counts at each loop pixel for biological replicates with mariner^68^ (version 1.2.1). This count matrix was used to identify differential loops with DESeq2^69^ (version 1.38.3) as previously described^8^. Loops with a median of 5 counts or less were removed before using a design of “∼rep + condition” to form a likelihood ratio test (LRT). Log_2_ fold-changes for each loop were computed with apeglm^70^ (version 1.20.0), comparing the proinflammatory to the resting condition. Significantly differential loops are those that have a FDR < 0.05 and log_2_ fold-change > 1.

#### ATAC-seq data processing and peak calling

Adapter sequences were removed using Trim Galore!^71^ (version 0.4.3). Alignment to the hg19 genome was then performed using BWA mem^72^ (version 0.7.17) and files were sorted and filtered for mitochondrial reads using Samtools^73^ (version 1.9). Duplicate reads were removed with PicardTools^74^ (version 2.10.3). The biological replicates for each condition were merged and indexed with Samtools, and the merged bam files were used to call peaks with the following MACS2 setting: “ -f BAM -q 0.01 -g hs --nomodel --shift 100 --extsize 200 --keep-dup all –B --SPMR” (version 2.1.2). A final peak list was generated by compiling all unique peak calls across both conditions (163,151 total peaks). The counts at each of these peaks for all biological replicates were extracted with bedtools^75^ multicov (version 2.30.0). Bigwig signals were generated from the merged conditions with deeptools^76^ (version 3.5.4).

#### ChIP-seq data processing and peak calling

Adapter sequences were removedusing Trim Galore!^71^ (version 0.403). Alignment to the hg19 genome was performed with BWA mem^72^ (version 0.7.17). Filtering and removal of mitochondrial reads was performed with Samtools^73^ (version 1.9) and duplicates were then removed with PicardTools^74^ (version 2.10.3). Biological replicates for each condition were merged and indexed with Samtools and the resulting bam files were used to call peaks with the following MACS2 setting: “-f BAM –q 0.01 -g hs --nomodel --shift 0 --extsize 200 --keep-dup all -B --SPMR” (version 2.1.2). A comprehensive peak list was generated by compiling unique peaks across both conditions (101,845 total peaks). Counts for each peak were extracted from individual replicates for each condition with bedtools^75^ multicov (version 2.30.0). Bigwig files were generated for signal visualization from the merged conditions with deeptools^76^ (version 3.5.4).

Differential ATAC-seq and ChIP-seq peak analysis

All differential peak analysis was performed with DESeq2^69^ (1.38.3). The resulting peak count matrix from ATAC-seq and ChIP-seq processing was read in with the function DESeqDataSetFromMatrix with a design of “∼rep + condition” and a reduced design of “rep” for a likelihood ratio test (LRT). A log_2_ fold-change between each condition was calculated with apeglm^70^ (1.20.0), and peaks were classified as significant if they had an FDR < 0.05 and an absolute log_2_ fold-change > 2.

#### TF motif enrichment

TF motif enrichment was performed on upregulated and downregulated ATAC peaks with the findMotifsGenome. pl function from HOMER (version 4.11). Peaks that were not significantly changing in accessibility between conditions were used as background peaks for both sets. The default parameters were used with “-size given”.

#### RNA-seq data processing and differential gene expression analysis

The quality of each fastq file was assessed with FastQC^77,78^ and MultiQC^77,78^ (FastQC version 0.11.5, MultiQC version 1.5). Adapters were then trimmed using Trim Galore!71 (version 0.4.3). Reads were quantified for hg19 with salmon^79^ (version 1.4.0) and alignment was performed with HISAT2^80^ (version 2.1.0). The resulting bam files were indexed with samtools^73^ (version 1.9). Samtools was used to merge the samples for each biological replicate, converted into bigwigs with deeptools^76^ (version 3.5.4) for visualization, and summarized as a txi with txImport^81^ (R version 4.2.2, tximport version 1.26.1). Differential gene expression analysis was performed with DESeq2^69^ (1.38.3). The txi file was imported into R using DESeqDataSetFromTXimport with a design of “∼rep + condition” and a reduced design of “rep” to form an LRT as described earlier for differential peak analysis. Shrunken log_2_ fold-changes were calculated using apeglm70 (version 1.20.0) for each gene comparing the proinflammatory condition to the resting condition. Genes were called significantly differential if they had an FDR < 0.05 and an absolute log_2_ fold-change > 2.

#### Heritability Enrichment Analysis

Heritability enrichment analysis was performed using stratified LD score regression (S-LDSC) analysis^82,83^ (version 1.0.0) for the following sets of peaks: resting ATAC, proinflammatory ATAC, resting H3K27ac, proinflammatory H3K27ac. LD scores were calculated from 1000 genomes European phase 3 bim file using the summary statistics from Jansen et al^30^.

#### TF motif enrichment and correlation with gene expression

Three motifs enriched in resting-specific or proinflammatory-specific macrophage ATAC peaks as described earlier were selected. Of the motifs selected, the top three most highly expressed genes in that category were selected to visualize.

#### Activity-By-Contact (ABC) model and differential ABC pair analysis

An adapted version of the ABC pipeline from Fulco et al 2019^39^ was used to identify enhancer-gene pairs. Briefly, one set of candidate enhancers was generated from the combination of resting and proinflammatory macrophage ATAC-seq data, and the top 150,000 strongest peaks were selected. Candidate peaks were resized to 500 bp in size from the summit, hg19 blacklisted were removed, and overlapping peaks were merged. Enhancer activity from H3K27ac ChIP and ATAC in resting and proinflammatory macrophages was extracted from each of the candidate regulatory elements and were normalized per replicate with variance stabilizing normalization. For each gene, ABC scores were calculated for all candidate elements in each condition using resting and proinflammatory macrophage Hi-C data in a 5 Mb window using mariner^68^. Enhancer-promoter pairs with an ABC score > 0.02 were called significant interactions. All ABC pairs were labeled with a specific enhancer-gene ID, and since the same enhancer list and gene list were used in each condition, this identifier was used to determine which ABC pairs were specifically valid in one context versus the other.

#### Gene Ontology (GO) enrichment analysis for ABC-anchored genes

Genes at the anchors of resting and activated-specific ABC pairs were subsetted for those that contained a differential gene and had a more strict ABC score > 0.05. These sets of genes (497 at proinflammatory ABC pairs and 561 at resting ABC pairs) were used as the input for gprofiler2^84^ (version 0.2.2) with all genes at ABC pairs as the background set (8,129 genes).

#### Comparison to Epigenomic Roadmap ATAC-seq data

ATAC-seq bigwigs from 53 cell types were downloaded from the Epigenomic Roadmap^41^ project. Each of the AD-associated genetic elements were categorized into 10 quantiles based on their MPRA activity levels (RNA/ DNA). For each element in each cell type, ATAC signal was extracted from the 150 bp region using the plotgardener^85^ readBigwig function (version 1.9.2). ATAC counts were first normalized across cell types and by cell type to control for differences in sequencing depth between cell types. Then quartile normalization was performed within each cell type.

#### TF motif disruption and enrichment analysis

The motifbreakR^45^ (version 2.12.3^45^) R package was used to determine what TF motifs our emVars may be disrupting. The motifbreakR function was used on each variant exactly as performed in McAfee et al^29^. The enrichment of all disrupted motifs was calculated with a Fisher’s exact test for the number of emVars vs the number of non-allelic and inactive variants that disrupt a given motif.

#### Comparison to public HEK MPRA experiments

The HEK293T MPRA data from Cooper et al^42^ was downloaded to compare with our THP-1 MPRA data. Benjamini-Hochberg corrections were applied to the 1,155 variants that were tested in both MPRA experiments, and variants with a new FDR of < 0.05 were called significant.

The variants from the HEK293T MPRA were grouped into 10 quantiles based on their activity (RNA/DNA) exactly as described above for our THP-1 MPRA. ATAC-seq counts were extracted from resting macrophage bigwigs for all variants using the plotgardener^85^ function readBigwig (version 1.9.2), and variants that were in the most active quantile in each experiment were compared to each other.

To compare emVars across studies, we identified emVars in the HEK293T MPRA by intersecting their MPRA-allelic variants and MPRA-active elements. The rsIDs for both MPRA were then overlapped with the microglia eQTL data^51^. IDE was determined by comparing the sign of eQTL beta to MPRA log_2_ fold-change after matching effect and non-effect alleles.

#### Comparison to cell type-specific H3K27ac data in the human brain

We analyzed H3K27 signatures of emVars in four major brain cell types from Nott et al^13^. For each emVar, we calculated the average H3K27ac signal from the 500 bp region flanking each variant from each cell type. The resulting H3K27ac signatures were visualized using the pheatmap package, clustering emVars by their similarities in cell type-specific H3K27ac signatures.

#### eQTL Integration with ABC to assign target genes

The microglia eQTL data from Kosoy et al^51^ was downloaded and overlapped with our variant rsIDs to connect to target genes (eGenes). Subsequently we subsetted for all significant variant-eGene pairs. This was done in parallel with overlapping the enhancer anchor of all ABC pairs across both conditions with all tested variants. We then subsetted emVars to identify their ABC-guided target genes. Using both methods, we linked 39 emVars to 76 target genes associated with AD risk (MPRA-AD risk genes). These 76 genes were used as an input for gprofiler2^84^ (version 0.2.2) in R to determine GO terms and KEGG pathways that are enriched.

#### Disease Associated Microglia Gene Signature Enrichment

RNA log_2_ fold-change from differential RNA-seq analysis between proinflammatory and resting macrophages was compared to genes either upregulated or downregulated in the disease associated microglia (DAM) gene signature from Hasselman et al^52^. The log_2_ fold-change between proinflammatory and resting macrophages intersecting with 416 genes upregulated in the DAM signature or 366 genes downregulated in the DAM signature was shown.

#### Comparison to single cell and single nuclear microglia RNA-seq data

Various datasets from Sun et al^38^ were downloaded and compared against MPRA-AD risk genes. Specifically, the microglial state markers for each of the 12 single-cell clusters were used for enrichment analysis versus genes that were expressed in our macrophages as background in a Fisher’s exact test. We also compared our MPRA-AD risk genes against DEGs describing the response of microglia to Aβ and Tau downloaded from Gazestani et al^24^. Similarly, we downloaded age-related DEGs from mouse microglia from Li et al 2023^53^ to compare against our MPRA-AD risk genes. We performed Fisher’s exact tests between MPRA-AD risk genes and each DEG set using all macrophage-expressed genes as background.

## Visualization

All locus plots using genomic signals were plotted with plotgardener^85^ (version 1.9.2). All other plots were made with ggplot2 (version 3.5.2).

## Data Availability

The THP-1 macrophage MPRA data will be available upon request. THP-1 macrophage Hi-C, ATAC-seq, H3K27ac ChIP-seq, and RNA-seq data are available under GEO: Superseries GSE201376.

## Data Availability

The THP-1 macrophage MPRA data is available under GEO: Series GSE273887. THP-1 macrophage Hi-C, ATAC-seq, H3K27ac ChIP-seq, and RNA-seq data are available under GEO: Superseries GSE201376.

## Code Availability

All code used to perform the analyses in this study are available at https://github.com/thewonlab/AD_MPRA

## Acknowledgement

We thank members of Won and Phanstiel labs for helpful discussions and comments about this paper. We thank Erika Deoudes for assistance in figure design and typesetting. This research was supported by the National Institute of Aging (R01AG066871, H.W., D.H.P.; R01NS128523, H.W.; F31AG84114, M.L.B.), the IGVF Consortium (UM1HG012003, H.W.), the National Institute of General Medical Sciences (5T32GM067553, S.L; 5T32GM135128, M.L.B., J.C.M.) and the BrightFocus Foundation (I.Y.Q-B fellowship 911831).

## Contributions

M.L.B., I.Y.Q-B., H.W., and D.H.P. designed the study. H.W. and J.C.M. designed the AD-MPRA library. J.C.M. generated the modified pLS-mP backbone. J.L.B. and M.P. generated the AD-MPRA library. I.Y.Q-B. and S.D. performed the MPRA experiments. S.D. worked on EMSA experimental design and Y.W. optimized and performed EMSA experiments. M.L.B. performed MPRA and genomic data processing, analysis, and integration. S.L., N.E.K., and M.L.B. performed eQTL overlap of emVars. H.W., D.H.P., M.L.B., I.Y.Q-B. co-wrote the manuscript, which was subsequently reviewed and edited by the rest of the authors.

