## Supplementary Information for "Deciphering the functional impact of Alzheimer’s Disease-associated variants in resting and proinflammatory immune cells"

#### Supplementary Tables

**Supplementary Table 1. Primer/oligo sequences used for MPRA library construction.**
Primer/oligo sequences used to generate the lentiviral MPRA backbone, to integrate AD-associated genetic elements into the lentiviral MPRA backbone, to generate MPRA libraries, and to perform EMSAs.

**Supplementary Table 2. RNA/DNA ratios for all elements tested.**
RNA/DNA ratios for all AD-associated genetic elements, positive controls, and negative controls for each biological replicate.

**Supplementary Table 3. MPRA summary statistics for transcriptional activity of AD-associated genetic elements.**
Transcriptional activity of AD-associated genetic elements over negative controls in both resting and proinflammatory macrophages. MPRA-active elements are specified.

**Supplementary Table 4. MPRA summary statistics for allelic regulatory activity of AD-associated variants.**
Allelic regulatory activity of AD-associated genetic variants in both resting and proinflammatory macrophages. MPRA-allelic variants and emVars are specified.

**Supplementary Table 5. ABC-defined enhancer-gene pairs**
ABC pairs in resting and proinflammatory macrophages with an ABC score > 0.02 in its given condition.

**Supplementary Table 6. TF motif disruption by emVars.**
Results from emVar motifbreakR analysis showing which emVars disrupt which TF motifs, pre-filtered for FDR < 0.05 and strong effects.

**Supplementary Table 7. emVar-gene relationship.**
Target genes of emVars from ABC and eQTL overlap.

### Supplementary Figures

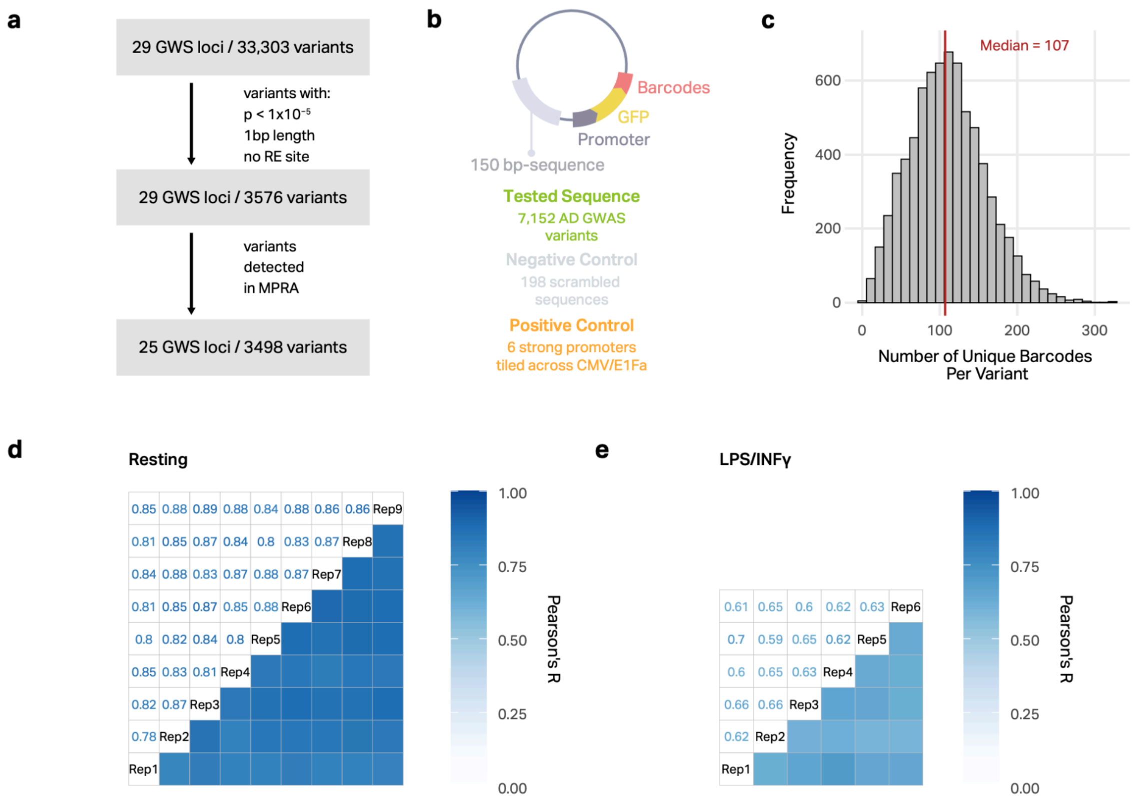


**Supplementary Figure 1. Overview of MPRA library design and quality control.**

**a,** Workflow of variant filtering from GWAS to final MPRA library design.

**b,** Schematic showing MPRA construct and the number of tested variants, scrambled negative controls, and positive controls.

**c,** Histogram showing the number of barcodes per variant detected in MPRA analysis.

**d-e,** Reproducibility analysis for all replicates from the resting macrophages (d) and LPS+INFγ-treated macrophages (e)

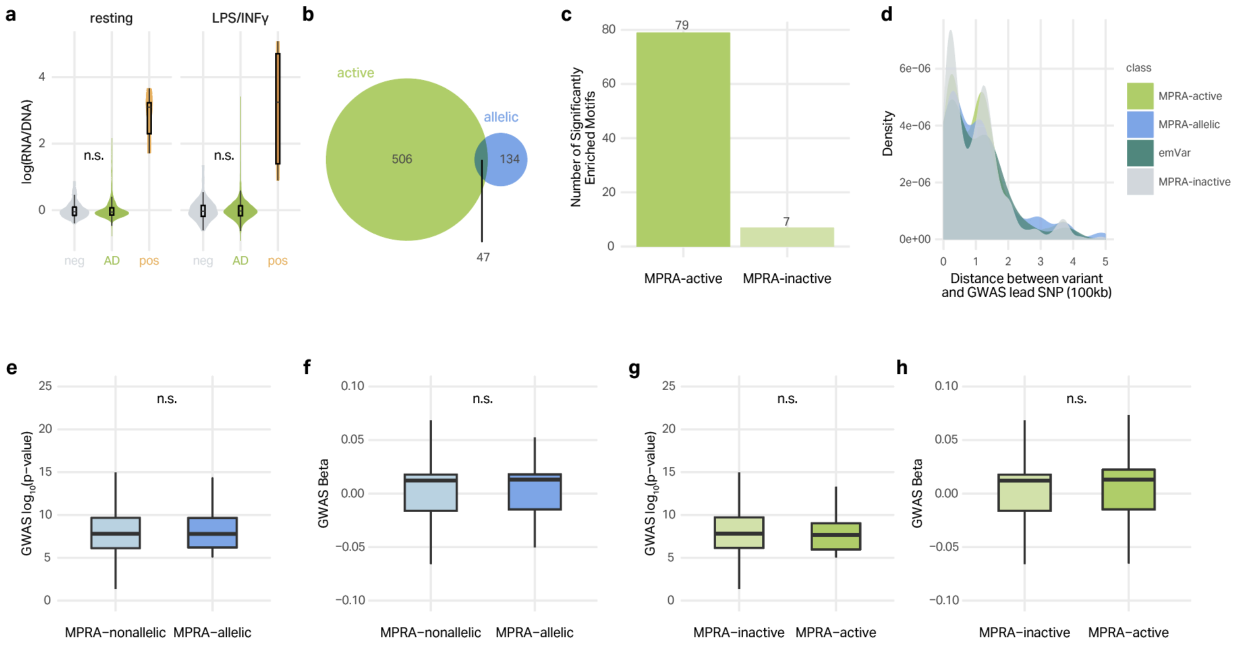


**Supplementary Figure 2. Characterization of MPRA-active elements and MPRA-allelic variants.**

**a,** Violin plots showing the activity of all AD tested elements before separating into MPRA-active and MPRA-inactive elements (AD), negative controls (neg), and positive controls (pose) for resting (left) and LPS+INFγ-treated (right) macrophages. Violins represent the distribution of the data, box plots show the median and 25th to 75th quartile with whiskers extending to the most extreme non-outliers. (n.s. represents $p > 0.05$ from two-sided Wilcoxon rank-sum test between negative controls and AD tested elements).

**b,** Venn diagram showing the overlap between MPRA-active elements and MPRA-allelic variants (emVars).

**c,** Barplot showing the number of significantly enriched TF motifs at MPRA-active and MPRA-inactive elements.

**d,** Density plot showing the distance between MPRA-active, MPRA-allelic, emVars, and MPRA-inactive/nonallelic elements and their respective lead GWAS variants. P-values calculated with a two-sided Wilcoxon rank-sum test (n.s. represents $p > 0.05$).

**e-f,** Box plots showing the difference in GWAS p-values (e) and beta values (f) for MPRA-allelic vs MPRA non-allelic variants. Box plots show the median and 25th to 75th quartiles with whiskers extending to the most extreme non-outliers (n.s. represents $p > 0.05$).

**g-h,** Boxplots showing the difference in GWAS p-values (g) and beta values (h) for MPRA-active vs MPRA-inactive elements. Box plots show the median and 25th to 75th quartiles with whiskers extending to the most extreme non-outliers (n.s. represents $p > 0.05$).

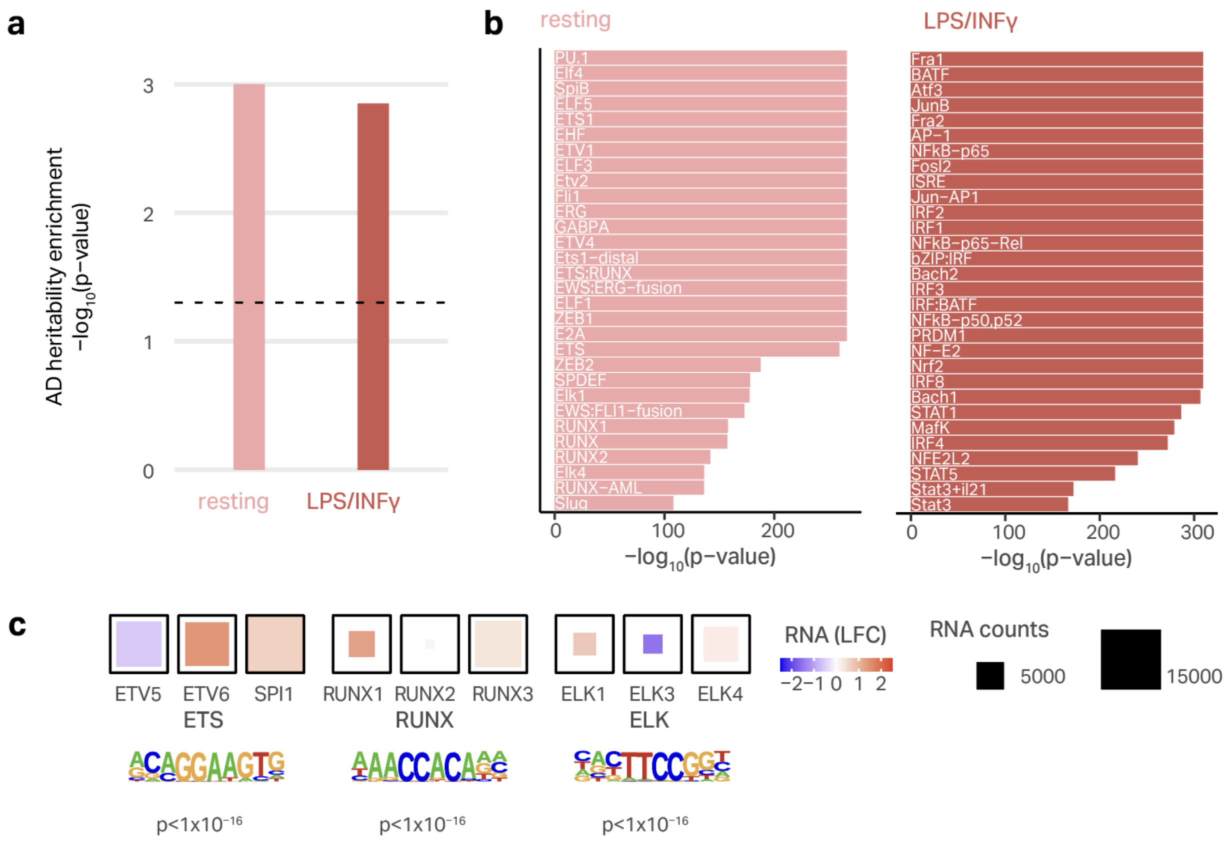


**Supplementary Figure 3. Additional epigenetic characterization of differential macrophage regulatory architecture.**

**a,** AD heritability enrichment in H3K27ac peaks from resting and LPS+INFγ-treated macrophages. P-values calculated by S-LDSC.

**b,** TF motifs enriched in resting-specific (left) and LPS+INFγ-specific (right) ATAC peaks.

**c,** TF motif enrichment for select motifs enriched in resting-specific ATAC peaks and the gene expression of the top three most highly expressed family members.

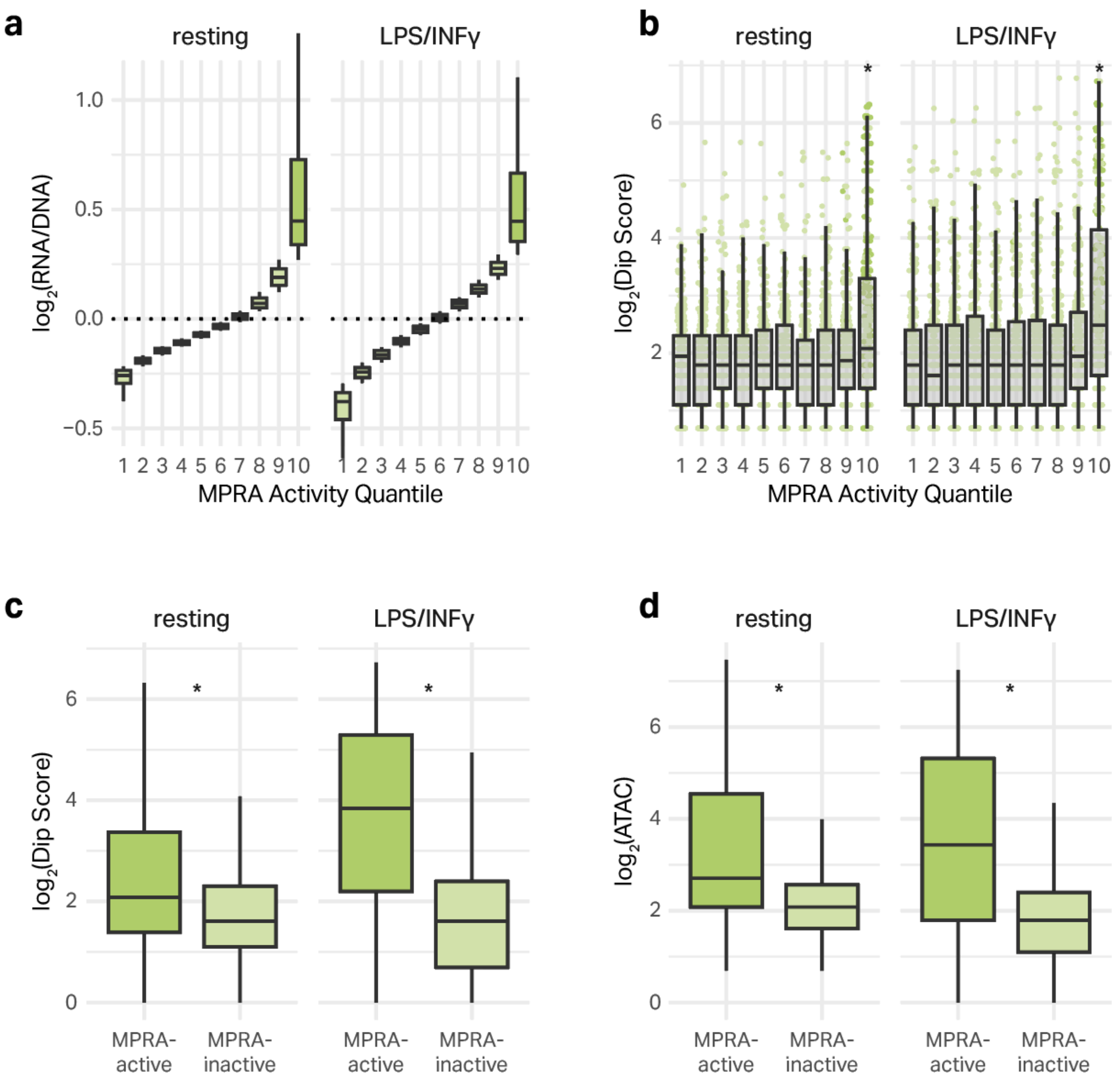


**Supplementary Figure 4. MPRA-active elements are more accessible and have higher H3K27ac signals than MPRA inactive elements**

**a,** Activity of MPRA elements in each of the ten quantiles (divided by MPRA-activity) in resting and LPS+INFγ-treated macrophages. Box plots show the median and IQR with whiskers extending to the most extreme non-outliers.

**b,** H3K27ac Dip Score for variants in each of the ten MPRA quantiles in resting and LPS+INFγ-treated macrophages. Box plots show the median and IQR with whiskers extending to the most extreme non-outliers. Asterisks represent two-sided Wilcoxon rank-sum test $p < 0.05$.

**c-d,** H3K27ac Dip Score (c) and ATAC counts (d) are higher in MPRA-active elements when compared to MPRA-inactive elements in resting and LPS+INFγ-treated macrophages. Box plots show the median and IQR with whiskers extending to the most extreme non-outliers. Asterisks represent two-sided Wilcoxon rank-sum test $p < 0.05$.

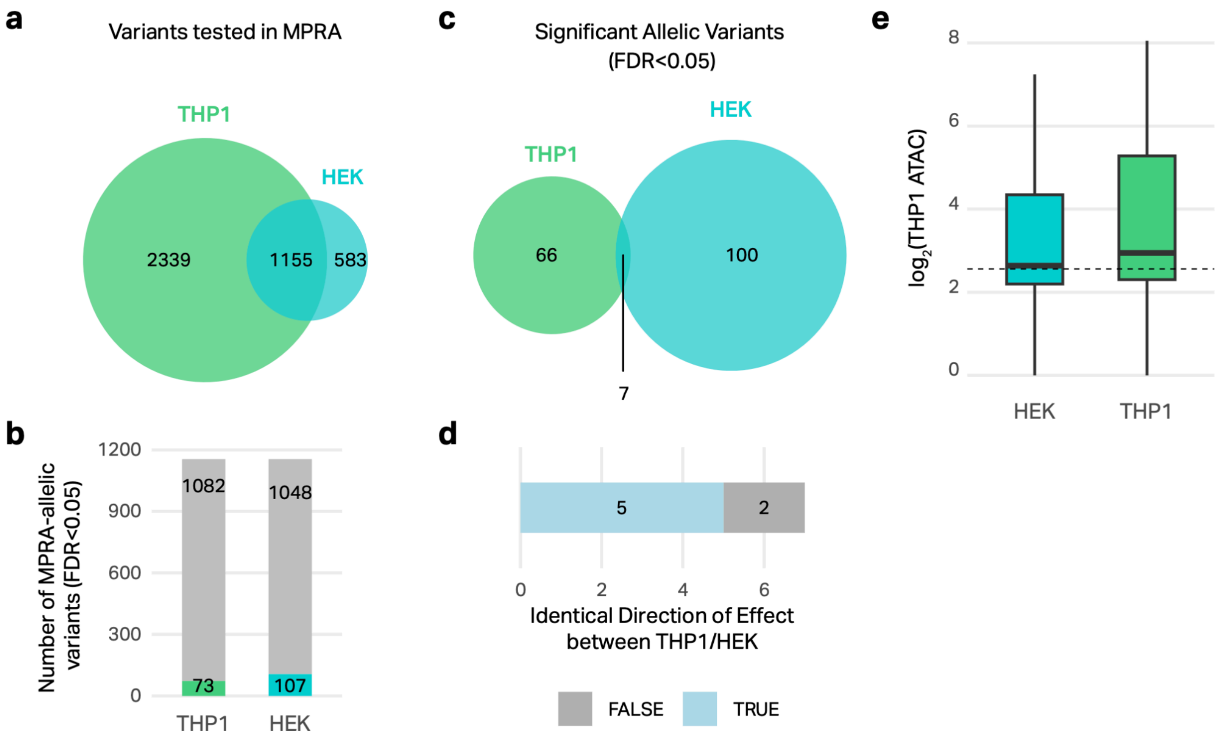


**Supplementary Figure 5. Comparison of THP-1 and HEK293 MPRA results.**
**a,** Venn diagram showing the variants tested in our THP-1 macrophage MPRA (THP1) compared to the AD-associated variants tested in the previously published HEK293T MPRA (HEK).
**b,** Barplot showing the number of tested variants that are MPRA-allelic in either THP-1 macrophages and HEK293T cells.
**c,** Venn diagram showing the overlap of MPRA-allelic variants in both THP-1 macrophages and HEK293T cells.
**d,** Barplot showing the direction of effect between the risk and protective alleles for the MPRA-allelic variants in both THP-1 macrophages and HEK293T cells.
**e,** ATAC-seq counts from THP-1 resting macrophages for HEK MPRA-active elements versus THP-1 MPRA-active elements. Box plots show the median and IQR with whiskers extending to the most extreme non-outliers (two-sided Wilcoxon rank-sum test, $p = 5.5 \times 10^{-4}$).

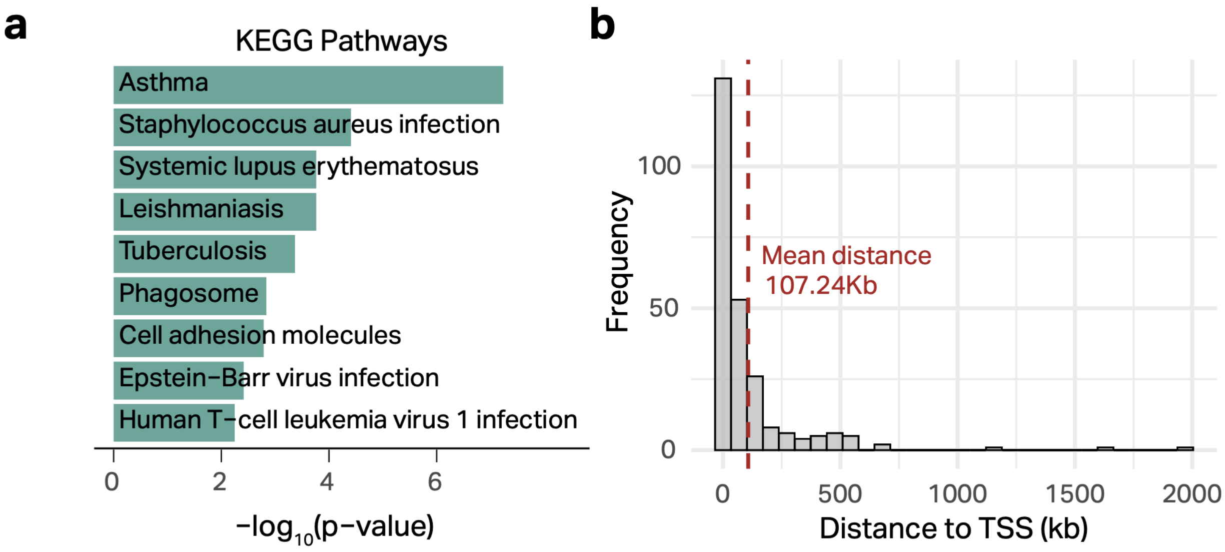


**Supplementary Figure 6. Additional characterization of emVar target genes.**
**a,** KEGG pathways enriched for the 76 emVar target genes.
**b,** Distances between emVars and their target gene TSS.
